# Contribution of maternal and fetal genetics to fetal weight throughout pregnancy and at birth

**DOI:** 10.64898/2026.10.04.26364695

**Authors:** Frances E Cates, Olivia Wootton, Jasmine X Kiley, Tabitha Wishlade, Ulla Sovio, Gordon C S Smith, Jasmine A Mack, D. Stephen Charnock-Jones, Hilary C Martin, Catherine E Aiken

## Abstract

**Background:** Both fetal and maternal genomes contribute to birthweight, but the timing and relative magnitude of their effects during gestation remain unclear. Polygenic risk scores (PRSs), constructed from genome-wide association studies (GWAS), capture the cumulative influence of common birthweight-associated variants to quantify genetic predisposition to fetal growth. We constructed fetal and maternal PRSs for birthweight in two UK birth cohorts, the Pregnancy Outcome Prediction Study (POPS) and Born in Bradford (BiB), to evaluate their associations with estimated fetal weight (EFW) throughout pregnancy and with birthweight at delivery. We also investigated whether PRSs could improve risk stratification of infants born below the 10th centile by distinguishing those with pathological intrauterine growth restriction (IUGR) from those who are constitutionally small-for-gestational-age (SGA).

**Methods:** In POPS and BiB, we used LDpred2-auto to estimate PRSs based on the largest available maternal and fetal GWAS for birthweight. We longitudinally assessed EFW associations with PRSs using linear mixed effects models in POPS and BiB. We evaluated birthweight associations with PRSs using linear models, where birthweight was adjusted for gestational age and fetal sex. We estimated R^2^ and conducted variance decomposition analyses for all models. Genetically-predicted birthweights were estimated from joint models of fetal and maternal PRSs plus ten genetic principal components (PCs). BiB analyses were conducted separately for genetically-inferred European (EUR) and South Asian (SAS) ancestry.

**Results:** While PRS_fetus_ was significantly associated with EFW at baseline (∼22 weeks gestation) across cohorts, PRS_mother_ was not. Both the interaction of PRS_fetus_ and PRS_mother_ with gestational age (GA) were significantly associated with EFW. The effect of PRS_mother_ on EFW increased more quickly across gestation than the effect of PRS_fetus_ in POPS, but this finding was not replicated in BiB. Jointly, fetal and maternal polygenic risk explained 4% of variance in EFW in POPS and 1% in BiB-EUR. In POPS, fetal and maternal PRSs showed independent contributions to EFW variance of 3% and 0.5%, respectively. Joint and univariable linear models showed significant positive associations of PRS_fetus_ and PRS_mother_ with observed birthweight. In POPS, PRS_fetus_ and PRS_mother_ accounted for ∼10% and ∼4% of variance in birthweight over covariates alone, respectively, which was replicated in BiB-EUR. Maternal and fetal genetic effects jointly explained 11% of birthweight variance in POPS, similar to BiB-EUR, at 12%; however, the variance explained was reduced by half in BiB-SAS, at 6%. Among POPS babies born <10th birthweight centile, the magnitude of the difference between observed and genetically-predicted birthweight was greater in the group judged clinically to be IUGR vs. SGA (p=0.008; point estimate=0.29, 95%CI: 0.12, 0.47).

**Conclusions:** Maternal and fetal PRSs jointly account for ∼11-12% of birthweight variance in European ancestry groups, but much less in a South Asian cohort. In POPS, the effect of birthweight-associated SNPs on estimated fetal weight increases more rapidly across pregnancy for maternal than for fetal genetic effects. Deviation from genetically-predicted birthweight may be a useful metric to help distinguish growth-restricted from constitutionally small babies.

1. **Tweetable statement:** Fetal and maternal genetics both influence fetal growth across gestation. Growth-restricted babies have greater differences between their genetically-predicted and observed birthweights compared to those who are constitutionally small.
2. **Short title:** Fetal and maternal genetic effects on fetal growth across gestation
3. **AJOG at a Glance:**

a. **Why was this study conducted?**

- We used a polygenic risk score (PRS) for birthweight to evaluate the effects of fetal and maternal genetics on estimated fetal weight (EFW) and birthweight across pregnancy and at birth.
- We also assessed how the difference between genetically-predicted and observed birthweight can help distinguish between small-for-gestational-age (SGA) babies and babies who experience intrauterine growth restriction (IUGR).
b. **What are the key findings?**

- Fetal PRS accounts for ∼11-12% of variance in birthweight, while maternal PRS accounts for ∼6% in two mother-child cohorts with European ancestry.
- In a South Asian ancestry cohort, the birthweight variance explained by the joint fetal-maternal PRS was approximately halved compared to European cohorts
- SGA babies show smaller differences between their predicted and observed birthweights compared to IUGR babies.
c. **What does this study add to what is already known?**

- This study separates the fetal and maternal genetic effects on EFW across gestation, and demonstrates that maternal genetic effects on fetal growth are more impactful at later gestations
- We demonstrate the need to include more diverse ancestry groups in large-scale genetic studies around pregnancy and birthweight to create more representative risk scores.
- Our findings suggest the potential use of deviation from genetically-predicted birthweight as a metric to differentiate growth-restricted (IUGR) from constitutionally small (SGA) babies.

## Introduction

Fetal growth is a dynamic developmental process shaped by fetal, placental, and maternal factors. It is an important determinant of perinatal outcomes and health across the life course^1–7^. Improving understanding of processes that shape fetal growth may provide insight into mechanisms underlying pregnancy complications and support identification of pregnancies at risk of adverse perinatal outcomes.

Birthweight reflects cumulative effects of fetal growth trajectories across gestation and timing of delivery. Epidemiological and genetic studies demonstrate that fetal and maternal factors contribute to birthweight variation^8–10^. SNP-based heritability for birthweight is 15-40%^10,11^, and genome-wide association studies (GWAS) show that birthweight is influenced by genetic variants carried by both mother and fetus^10^. Fetal genetic effects may capture influences acting through the fetus or placenta, whereas maternal genetic effects may act indirectly, shaping the intrauterine environment. Although both contribute, fetal genetic factors likely explain more of the total birthweight variance^10^.

However, using birthweight as a stand-alone metric provides limited insight into when influences on fetal growth emerge, how they vary across pregnancy, or whether they affect fetal size, gestational duration, or both^12^. The relative influence of fetal and maternal genetic contributions may vary across gestation^13^. As fetal nutrient demand increases, placental transport capacity and maternal nutrient supply are likely to become increasingly relevant to fetal size^14,15^. However, the lack of cohorts with maternal and fetal genetic data alongside universal serial ultrasound measures has prevented systematic understanding of how genetics shape fetal growth trajectories.

A key reason for seeking further insight into genetic influences on fetal growth is that it remains complex to distinguish between small (<10th centile birthweight) babies that fail to meet their genetic growth potential (intrauterine growth restriction, or IUGR) and constitutionally small, healthy babies (small-for-gestational-age, or SGA). These groups exhibit different risk profiles for immediate and long-term adverse outcomes^16–18^. Existing clinical approaches to identifying IUGR^19–22^ are heterogeneous and fail to accurately classify all cases. We assess whether the difference between observed and genetically-predicted birthweight can be useful to distinguish IUGR from SGA babies.

Further, birthweight distributions differ across ethnic groups^23^, yet most genetic studies of birthweight are conducted in individuals of genetically-inferred European ancestry. Previous evidence suggests that while common variant effects acting through the fetus may be consistent across populations^10^, there may be differing associations between maternal PRS and birthweight across ethnic groups^24^. Using data from two ethnically, socio-economically, and geographically distinct UK birth cohorts, the Pregnancy Outcomes Prediction Study (POPS) and Born in Bradford (BiB), we investigated the extent to which maternal and fetal polygenic risk explain variation in (i) estimated fetal weight (EFW) across pregnancy, (ii) birthweight, and (iii) the likelihood of IUGR at birth.

## Methods

### 1. Cohort Overview

#### a. Pregnancy Outcome Prediction Study (POPS)

POPS is a prospective study of unselected nulliparous women at Rosie Hospital in Cambridge, United Kingdom. Women with a singleton pregnancy were enrolled from January 2008-July 2012 (n=4,212)^25,26^. All participants underwent standardized, serial ultrasound scans at ∼12, ∼20, ∼28, and ∼36 weeks gestation^26^ (Table S1, Supplemental File 1). Within the analytic sample, all POPS participants self-reported their ethnicity as White British; 25% of participants were in the most deprived socioeconomic quartile (Table S2).

#### b. Born in Bradford (BiB)

BiB is a multi-ethnic, longitudinal birth cohort (n=12,453) recruited between 2007-2011^27^. 52% of the cohort reported South Asian ethnicity; 45% of European ethnicity and 68% of South Asian ethnicity participants were from neighborhoods in the most deprived socioeconomic quintile (Table S2). BiB did not include universal serial ultrasound scans, however, many participants had clinically-indicated ultrasound scans after their ∼20 week scan (Table S1), the results of which were extracted from maternal health records.

### 2. Preparation of the phenotype data

Observed birthweight was converted to age- and sex-adjusted birthweight centiles using the UK 1990 growth reference data^28^. In POPS, n=2543 babies were included in birthweight analyses, compared to n=2075 in BiB-EUR and n=2210 in BiB-SAS (Table S1).

In POPS, gestational age was derived from crown-rump length (CRL) measured by ultrasound (<14 weeks of gestation)^20^. In BiB, gestational age was determined based on last menstrual period (LMP) and confirmed by ultrasound scan^29^. Estimated fetal weight (EFW) was calculated using the Hadlock III formula^30^. In POPS, gestational age (GA)-adjusted z-scores for EFW were estimated using the cohort-specific reference distribution based on all four biometric measurements: abdominal circumference, femur length, head circumference, and biparietal diameter^31^. Data were available for n=2531 children with scans at 18-22 weeks, n=2515 at 26-30 weeks, and n=2396 at 34-38 weeks (Table S1). In BiB, gestational age-adjusted z-scores for EFW were calculated using INTERGROWTH-21st standards^32^. In BiB-EUR, data were available for n=1358 children with scans at ∼20 weeks, compared to n=1435 in BiB-SAS; data were also extracted for a second, later scan for n=728 children in BiB-EUR and n=749 in BiB-SAS (Table S1). We excluded multiple pregnancies and pregnancies for which a congenital anomaly was reported.

### 3. Processing and preparation of the genotype data

We describe the procedures for genotyping, imputation, kinship analysis, and principal component analysis in POPS and BiB in Supplemental File 2.

### 4. Construction of polygenic risk scores

We constructed a PRS for birthweight using the largest maternal and fetal GWAS for birthweight by deCODE, which was conducted in individuals of European ancestry^33,34^. SNP weights were estimated using LDpred2-auto with 1,444,196 HapMap3+ variants^35,36^ and a precomputed LD reference panel provided by the LDpred2 authors.

SNP weights were applied separately in BiB and POPS to calculate fetal and maternal PRSs for birthweight. We used the fetal genotype to estimate a PRS for fetal birthweight (PRS_fetus_), and the mother’s genotypes to estimate a PRS for offspring birthweight (PRS_mother_), with each PRS standardized (mean = 0, SD = 1) within each analytic cohort.

### 5. Statistical analysis

#### a. Estimating genetic associations with Estimated Fetal Weight

EFW_z_ represents the z-score for estimated fetal weight, standardized for gestational age. Where serial ultrasound data was available, we fit linear mixed effects models using continuous GA, a full description of which is in Table 1A. Covariates included in adjusted models were: maternal age at first appointment, maternal height, paternal height, and the first 10 genetic principal components (PCs). In BiB, maternal and paternal height were excluded due to high missingness.

**Table 1A.** Longitudinal and cross-sectional models of estimated fetal weight in POPs and BiB. This table describes the linear mixed effects models (1-5) used in POPs and BiB and the linear models (6-10) used in BiB to assess contributions of polygenic risk scores to estimated fetal weight (EFW) across gestation. Covariates included maternal age at first appointment, maternal height, paternal height, and the first ten genetic principal components (PCs). GA is modelled as a continuous variable centered on 22 weeks. 1 | represents the per-pregnancy random intercept. As a robustness analysis in POPS, we evaluated timepoint as a categorical variable using model 4, where we characterized 3 gestational ages corresponding to POPS scans at 20, 28, and 36 weeks. For models 1-5 in POPS, we calculated marginal R^2^, conditional R^2^, incremental (Δ) R^2^, and AIC values to assess variance in EFW explained by (1) covariates only, (2) PRS_fetus_, (3) PRS_mother_, (4) PRS_fetus_ + PRS_mother_, and (5) PRS_fetus_ PRS_mother_. Marginal R^2^ represents the variance explained by fixed effects only, while conditional R^2^ represents the variance explained by both fixed and random effects. Incremental (Δ) R^2^ is defined as the difference in marginal R^2^ between each model and the baseline (covariates-only) model (1), quantifying the additional proportion of variance in EFW explained by fixed effects beyond baseline, holding the random-effects structure constant. We determined the independent effects of maternal and fetal genetics on EFW_z_ by calculating the difference between the joint model (4) and the univariable PRS_fetus_ (2) and PRS_mother_ (3) models, respectively. For cross-sectional models 6-10 in BiB, we calculated R^2^, incremental (Δ) R^2^, and AIC values to assess EFW explained by (6) baseline covariates, (7) PRS_fetus_, (8) PRS_mother_, (9) PRS_fetus_ + PRS_mother_, and (10) PRS_fetus_ PRS_mother_. In the covariate-only model, BiB included maternal age but not maternal and paternal height due to high missingness.

|  |  |
| --- | --- |
| **Model 1** | $EFW_z \sim \text{covariates} + GA + (1\vert pregnancy)$ |
| **Model 2** | $EFW_z \sim PRS_{fetus} + \text{covariates} + GA + (GA \times PRS_{fetus}) + (1\vert pregnancy)$ |
| **Model 3** | $EFW_z \sim PRS_{mother} + \text{covariates} + GA + (GA \times PRS_{mother}) + (1\vert pregnancy)$ |
| **Model 4** | $EFW_z \sim PRS_{fetus} + PRS_{mother} + \text{covariates} + GA + (GA \times PRS_{fetus}) + (GA \times PRS_{mother}) + (1\vert pregnancy)$ |
| **Model 5** | $EFW_z \sim PRS_{fetus} \times PRS_{mother} + \text{covariates} + GA + (GA \times PRS_{fetus}) + (GA \times PRS_{mother}) + (1\vert pregnancy)$ |
| **Model 6** | $EFW_z \sim \text{covariates}$ |
| **Model 7** | $EFW_z \sim PRS_{fetus} + \text{covariates}$ |
| **Model 8** | $EFW_z \sim PRS_{mother} + \text{covariates}$ |
| **Model 9** | $EFW_z \sim PRS_{fetus} + PRS_{mother} + \text{covariates}$ |
| **Model 10** | $EFW_z \sim PRS_{fetus} \times PRS_{mother} + \text{covariates}$ |

In linear mixed effects models, fixed effects represent the individual PRS effects centered at 22 weeks, while the interaction term (PRS GA) represents changes in EFW_z_ over time. We determined independent effects of maternal and fetal genetics on EFW_z_ by calculating the difference between joint and univariable modelling of PRS_fetus_ and PRS_mother_. We assessed the interaction effect of PRS_fetus_ and PRS_mother_ using likelihood ratio testing.

#### b. Estimating genetic associations with birthweight

We tested for associations of birthweight with PRS_fetus_ and PRS_mother_ using a series of five linear models, sequentially including fetal and maternal PRSs to assess their independent and joint effects (Table 1B), including an interaction term (PRS_fetus_ PRS_mother_) to test whether PRS_mother_ modifies the association with PRS_fetus_.

**Table 1B.** Linear models for assessing birthweight associations in POPs and BiB. This table describes models used in POPs and BiB to assess associations of polygenic risk scores with birthweight. In these models, BW_z_ represents birthweight standardised for gestational age and sex. Covariates include maternal age, maternal height, and paternal height in POPs and maternal age in BiB. We calculated R^2^, incremental (Δ) R^2^, and AIC values to assess birthweight explained by (11) baseline covariates, (12) PRS_fetus_, (13) PRS_mother_, (14) PRS_fetus_ + PRS_mother_, and (15) PRS_fetus_ PRS_mother_. Incremental R^2^ values represent the additional variance in BW_z_ relative to that explained by the covariates-only model (11). We assessed the independent contributions of maternal and fetal genetics by calculating the difference between the joint model (14) and the univariable PRS_fetus_ (12) and PRS_mother_ (13) models, respectively.

|  |  |
| --- | --- |
| **Model 11** | $BW_z \sim \text{covariates}$ |
| **Model 12** | $BW_z \sim PRS_{\text{fetus}} + \text{covariates}$ |
| **Model 13** | $BW_z \sim PRS_{\text{mother}} + \text{covariates}$ |
| **Model 14** | $BW_z \sim PRS_{\text{fetus}} + PRS_{\text{mother}} + \text{covariates}$ |
| **Model 15** | $BW_z \sim PRS_{\text{fetus}} \times PRS_{\text{mother}} + \text{covariates}$ |

#### c. Construction of term for deviation from genetically-predicted birthweight

We used five-fold cross-validation to estimate a deviation term (D) representing deviation of observed birthweight from genetically-predicted birthweight in each cohort. In training folds, we fit the model *BW*_z_∼PRS_fetus +_PRS_mother_ +*PC*1…10. We then predicted *BW*_z_ (the genetically-predicted value for BW_z_) in the held-out fold and estimated D using the equation BW_z_, where D represents the difference between the observed and genetically-predicted birthweight z-score.

We compared polygenic risk across babies born <10th centile in POPS only, due to data availability. IUGR was defined as birthweight <10^th^ centile with clinical evidence of growth restriction (defined in Table S3)^37^. SGA was defined as birthweight<10^th^ centile without clinical evidence of growth restriction. Other babies were categorized as appropriate for gestational age (AGA; birthweight ≥10^th^ and ≤90^th^ centile) or large for gestational age (LGA; birthweight >90^th^ percentile).

#### d. Correction for multiple testing

We corrected for multiple testing using the Benjamini-Hochberg procedure to control the false discovery rate (FDR) at 5%. In EFW models (1-10), we applied the correction to PRS_fetus_, PRS_mother_, PRS x GA, and GA. In birthweight models (11-15), we applied the correction to p-values for PRS_fetus_, PRS_mother_, and PRS_fetus_ x PRS_mother_. All reported q-values in the results are FDR corrected per the Benjamini-Hochberg method.

## Results

### 1. Influence of fetal and maternal polygenic risk scores on EFW across gestation

Analysis in POPS showed that the strength of association between EFW_z_ and both fetal and maternal PRS increased as gestation progressed (Figure 1). When only fetal genetics were considered, both PRS_fetus_ (q=1.2e-18) and the interaction between PRS_fetus_ and gestational age (q=1.1e-04) were associated with EFW_z_ (Table S4A). Considering only maternal genetics, PRS_mother_ was not significantly associated with EFW_z_ at baseline (∼22 weeks; q = 0.121), although there was a significant interaction between PRS_mother_ and increasing gestational age (q=3.4e-11) (Table S4B).

**Figure 1:**
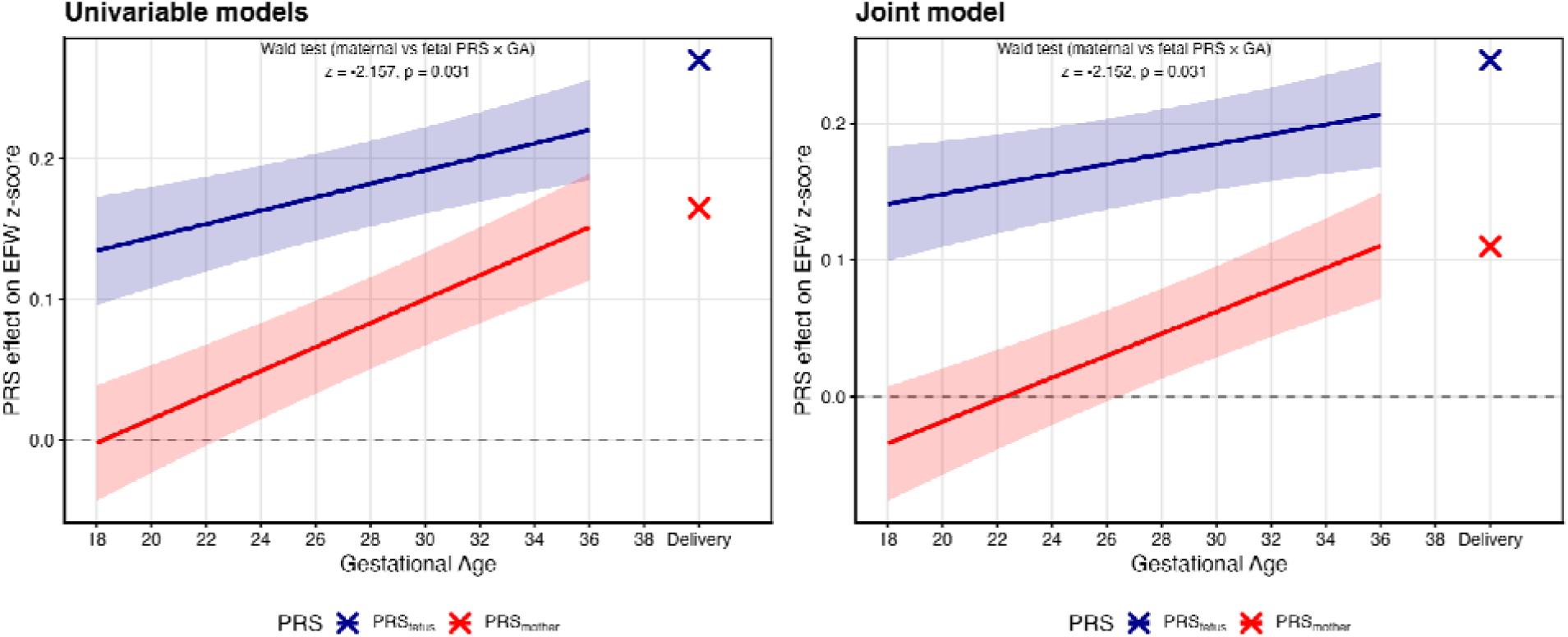
Birthweight PRS associations with EFW_z_ across gestation from univariable and joint linear mixed-effects models (POPS). Lines show the estimated effect of each birthweight PRS on EFW at each gestational week derived from linear mixed-effects models including a PRS × gestational age interaction term. The left panel shows estimates from univariable models in which PRS_fetus_ and PRS_mother_ were fitted in separate mixed-effects models. The right panel shows trajectories from a joint mixed-effects model in which both PRSs were included simultaneously. Ribbons show 95% confidence intervals. The cross at “Delivery” shows the corresponding PRS association with observed birthweight.

When fetal and maternal genetics were assessed in the same model (model 4), PRS_fetus_ (q=2.1e-16) but not PRS_mother_ (q=0.915) was significantly associated with EFW_z_ at ∼22 weeks. Both the interaction of PRS_mother_ (q=1.6e-09) and PRS_fetus_ (q=0.008) with GA were significant (Table 2; Table S4C), including after adjustment for socioeconomic status and other covariates (Table S4D). The effect of PRS_mother_ on EFW_z_ increased significantly more quickly across gestation than that of fetal PRS (contrast estimate = -0.004, Z = -2.15, p = 0.031; Table S5).

**Table 2.**
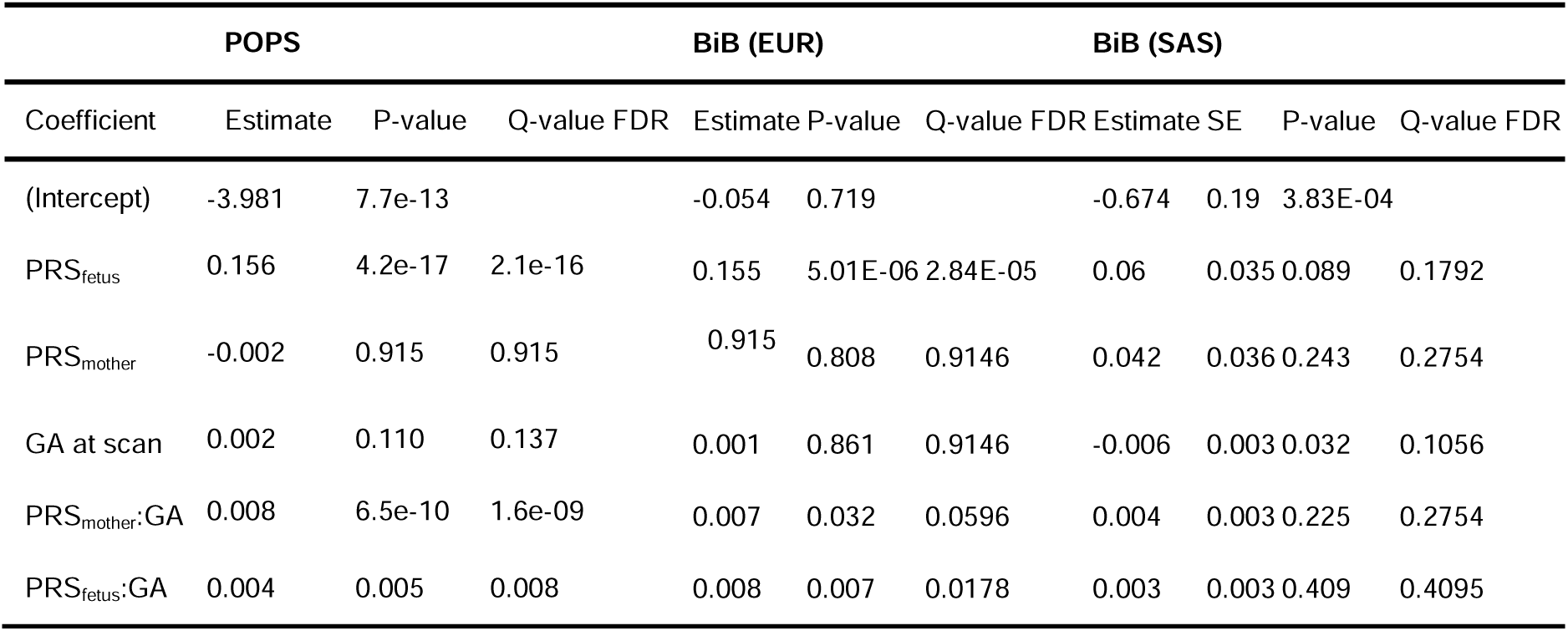
Joint model (4) of longitudinal associations between PRS_fetus_ and PRS_mother_ and EFW_z_ with continuous GA in POPS and BiB. Gestational age (GA) was modeled as a continuous variable to assess changes across gestation in the associations of PRS_mother_ and PRS_fetus_ with EFW_z_ using model 4. Regression coefficients represent fixed effects from the joint model, including PRS x GA interaction terms. Estimates are presented with Wald test p-values and false discovery rate (FDR) q-values. FDR correction was applied using the Benjamini-Hochberg method. Here we show only predictors of interest but the full model is reported in Table S2C.

In POPS, PRS_mother_ and PRS_fetus_ accounted for substantially more variance in EFW_z_ than covariates alone (Figure S1). Fetal and maternal genetic effects jointly explained 3.95% of variance in EFW_z_ over the covariates-only model (Table S6A). Variance decomposition showed independent contributions of PRS_fetus_ and PRS_mother_ of 2.92% and 0.45%, respectively, to EFW_z_ (Table S6B). Including an interaction term between PRS_fetus_ and PRS_mother_ did not significantly improve model fit compared to the joint model (p=0.129; Table S6C).

We tested whether the associations of maternal and fetal PRSs with EFW at ∼22 weeks in POPS were consistent with cross-sectional analysis in BiB. In BiB-EUR and BiB-SAS, we observed an association between PRS_fetus_ and EFW at ∼22 weeks (BiB-EUR, q=8.83e-05; BiB-SAS, q=0.024) (Table S7A). PRS_fetus_ explained 1.3% and 0.4% of the variance in EFW in BiB-EUR and BiB-SAS, respectively (Figure S2A&B), compared to 3.51% in POPS (Figure S1).

In BiB-EUR, like POPS, there was no significant association between PRS_mother_ and EFW at ∼22 weeks (q=0.688); however, a positive association was observed in BiB-SAS (q = 0.006; Table S7B). These trends in PRS_mother_ persisted in the joint model, as did the association of PRS_fetus_ with EFW_z_ (Table S7C). Jointly, PRS_fetus_ and PRS_mother_ accounted for 1.28% of variance in EFW_z_ for BiB-EUR and 0.94% for BiB-SAS (Table S8A). Variance decomposition in BiB-EUR showed that PRS_fetus_ and PRS_mother_ independently accounted for 1.24% and 0.01% of variance in EFW_z_, respectively (Table S8B).

In the subset of BiB-EUR participants with serial EFW measurements (n=728), both the fetal and maternal birthweight PRS associations with EFW strengthened across gestation (Figure 2A; Table 2). When fitted alone, PRS_fetus_ was significantly associated with EFW ∼22 weeks (q=2.84e-05); this association increased with gestational age (q=0.013) (Table S4A). In the joint and univariable models, PRS_mother_ showed no significant association with EFW at ∼22 weeks, but there was a significant interaction with increasing gestational age, reflecting an effect of PRS_mother_ on EFW that emerges later in gestation (univariable, q=0.004; joint, q=0.018) (Table S4B; Table 4C). There was no significant difference between the PRS_fetus_ × GA and PRS_mother_ × GA interaction slopes in either BiB ancestry group (BiB-EUR, p=0.74; BiB-SAS: p=0.82) (Table S5).

**Figures 2A&B.**
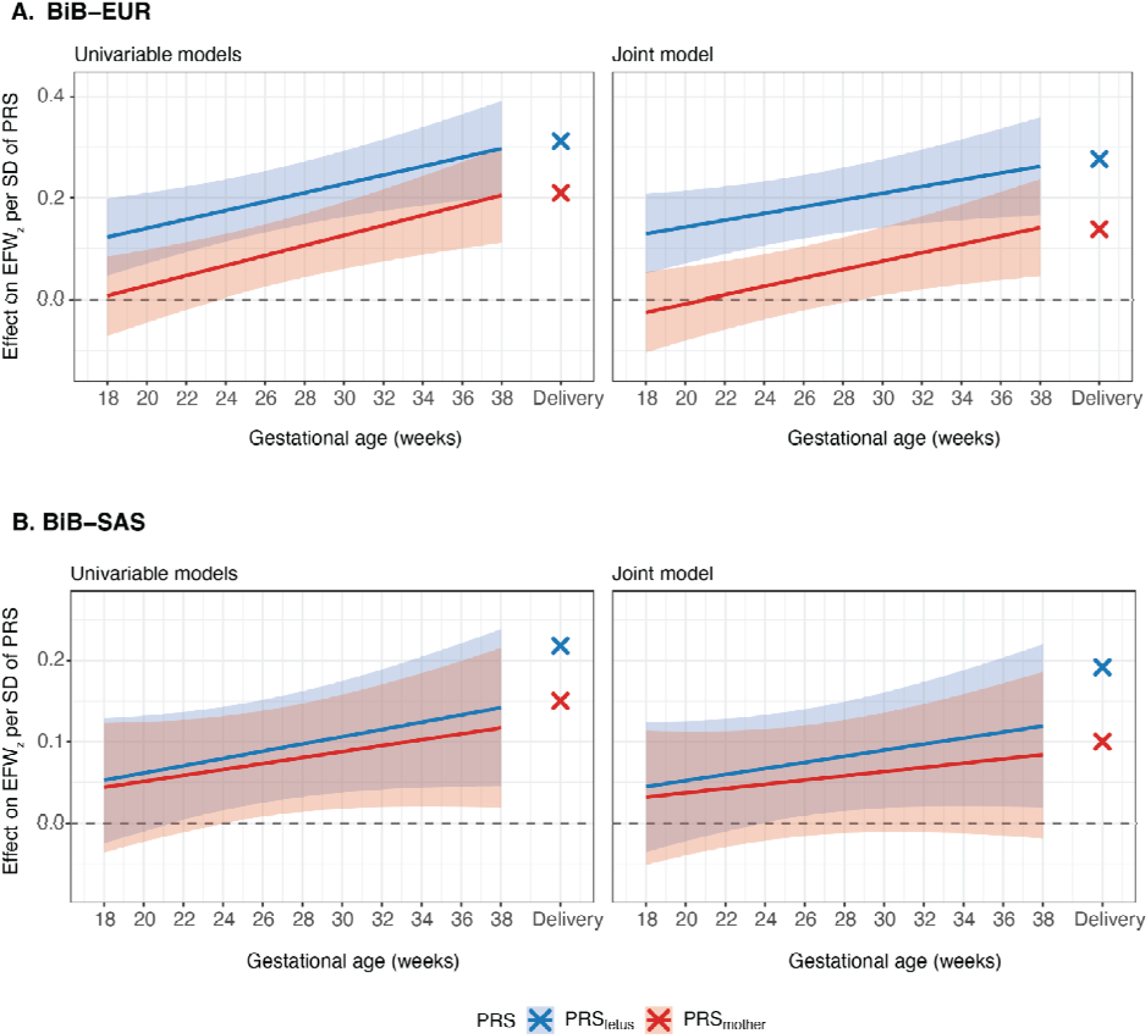
PRS associations with EFW across gestation from univariable and joint linear mixed-effects models in BiB. Lines show the estimated effect of each birthweight PRS on EFW at each gestational week derived from linear mixed-effects models including a PRS × gestational age interaction term. The left panel shows estimates from univariable models in which PRS_fetus_ and PRS_mother_ were fitted in separate mixed-effects models. The right panel shows trajectories from a joint mixed-effects model in which both PRS were included simultaneously. Ribbons show 95% confidence intervals. The cross at “Delivery” shows the corresponding PRS association with observed birthweight. EUR: genetically-inferred European ancestry (N=728); BiB-SAS: genetically-inferred South Asian ancestry (N=749).

### 2. Influence of fetal and maternal polygenic risk scores on birthweight

Across cohorts, we observed a significant positive association between PRS_fetus_ and PRS_mother_ and birthweight in univariable (models 12-13) (Table S9A&B) and joint models (models 14-15) (Table 3; Table S9C&D). Variance in birthweight explained by fetal and maternal PRSs was substantially higher in both of the European cohorts (POPS, BiB-EUR) compared to BiB-SAS (Figure S3A, S3B, S3C). In POPS, PRS_fetus_ and PRS_mother_ jointly accounted for 11.18% of the variance in birthweight over covariates alone (Figure 3A; Table S10A), comparable to 12.23% in BiB-EUR (Figure 3B; Table S10A). The corresponding variance explained in BiB-SAS was lower at 6.29% (Figure 3C). Variance decomposition confirmed an independent contribution of each PRS to birthweight variance across cohorts, with lower explanatory power in BiB-SAS (Table S10B).

**Table 3.** Associations between fetal and maternal polygenic risk scores (PRS) and observed birthweight z-scores across POPS, BiB-EUR, and BiB-SAS (Model 14). We assessed the cross-sectional associations of PRS_mother_ and PRS_fetus_ using the joint model (Model 14) across cohorts. Estimates are presented with Wald test p-values and false discovery rate (FDR) q-values. FDR correction was applied using the Benjamini-Hochberg method to generate q-values. Adjustments in POPS included maternal age at booking, maternal height, paternal height, PCs 1-10, while BiB adjusted for maternal age at booking and PCs 1-10. Here we show only predictors of interest but the full model is reported in Table S8D.

| Coefficient | POPS |  |  | BiB (EUR) |  |  | BiB (SAS) |  |  |
| --- | --- | --- | --- | --- | --- | --- | --- | --- | --- |
|  | Beta | P-value | Q-value FDR | Beta | P-value | Q-value FDR | Beta | P-value | Q-value FDR |
| (Intercept) | -4.166 | 3.31e-14 |  | -0.706 | 1.15e-13 |  | -1.157 | 2.08e-26 |  |
| PRS<sub>fetus</sub> | 0.246 | 3.67e-48 | 1.14e-47 | 0.277 | 1.25e-40 | 3.51e-40 | 0.192 | 1.61e-21 | 3.75e-21 |
| PRS<sub>mother</sub> | 0.11 | 3.58e-11 | 5.02e-11 | 0.138 | 1.65e-11 | 1.93e-11 | 0.1 | 5.05e-07 | 7.08e-07 |

**Figures 3A,B,&C.**
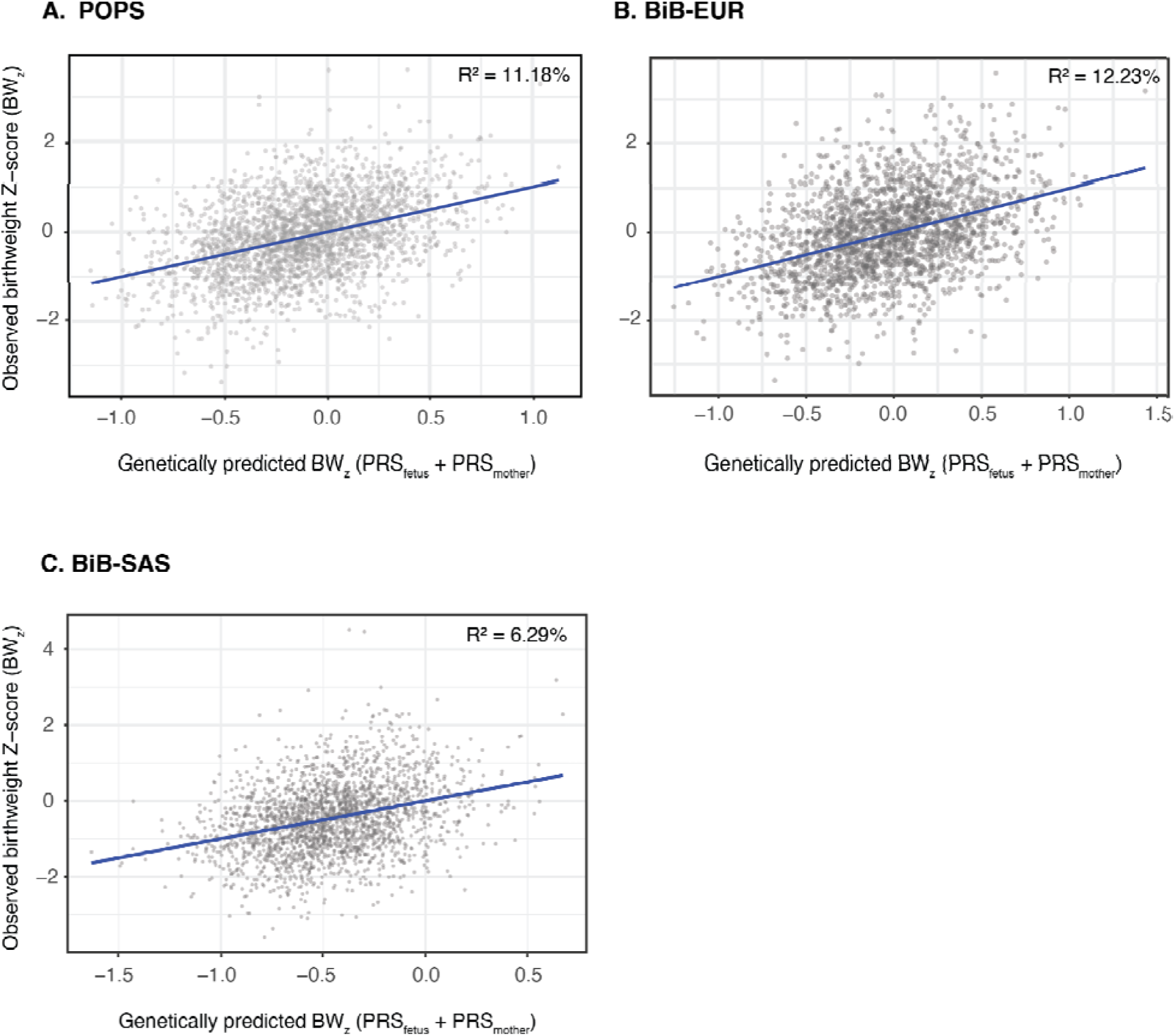
The association between observed birthweight Z-score (BW_z_) and genetically-predicted BW_z_. Predicted values were fitted from a linear model regressing BW_z_ on PRS_fetus_,PRS_mother_, and covariates in each cohort (model 14, Methods). R^2^ is the incremental variance explained by PRS_fetus_ and PRS_mother_ over covariates alone. BiB-EUR: genetically-inferred European ancestry participants from Born in Bradford (N=2,075); BiB-SAS: genetically-inferred South Asian ancestry from Born in Bradford (N=2,210); POPS: Pregnancy outcomes prediction study (N=2,537).

### 3. Deviation from genetically-predicted birthweight in POPS

The average birthweight genetic risk score for mother and fetus differed significantly across observed birthweight groups in POPS (AGA, LGA, and SGA/IUGR; Figure S4). Babies born AGA and LGA had higher mean PRSs than babies born <10^th^ birthweight centile (Table S11A); however, there was no significant difference in mean PRS of mother or fetus between the IUGR and SGA groups (Table S11B&C). LGA babies tended to be larger than their genetically-predicted size, whereas both SGA and IUGR babies tended to be smaller (Table S12). Although PRS alone did not differentiate between IUGR and SGA groups, IUGR babies had a significantly more negative mean difference between their predicted and observed birthweights than SGA babies (p=0.008, Table 4).

**Table 4.** Pairwise differences in deviation from genetically predicted birthweight across fetal growth classifications. Pairwise t-tests comparing mean deviation between fetal growth categories (IUGR, SGA, AGA, and LGA). Deviation represents the difference between the observed and genetically-predicted birthweight z-score, where *D* = *BW_z_ BW͡_z_*. For each comparison, we report the mean difference in D (second group as reference), 95% confidence intervals, Cohen’s d effect size, and associated p-values. Positive values indicate higher deviation in the first-listed group relative to the reference group, while negative values indicate lower deviation. We applied the Bonferroni correction to P-values.

| Comparison | Mean diff in mean D | CI | Cohen's d | P-value |
| --- | --- | --- | --- | --- |
| IUGR vs AGA | 1.67 | [1.51, 1.84] | 2.73 | p = 2.1e-26 |
| SGA vs AGA | 1.38 | [1.3, 1.46] | 2.27 | p = 1.8e-80 |
| LGA vs AGA | -1.54 | [-1.65, -1.44] | -2.52 | p = 3.4e-57 |
| IUGR vs SGA | 0.29 | [0.12, 0.47] | 0.57 | p = 0.008 |
| IUGR vs LGA | 3.22 | [3.03, 3.4] | 5.75 | p = 4.3e-54 |
| SGA vs LGA | 2.92 | [2.79, 3.05] | 5.56 | p = 1.6e-114 |

## Discussion

### a. Principal Findings

Our findings outline the relative fetal and maternal contributions to fetal weight throughout gestation and at birth. We demonstrate that fetal PRS is associated with EFW measurements from ∼22 weeks of pregnancy onwards. In the POPS cohort, using universal serial measurements, the strength of the association between both fetal and maternal genetics and EFW increased as pregnancy progressed. We show that maternal genetic effects on EFW increase over gestation to a greater extent than corresponding fetal genetic effects in POPS. This effect was not replicable in BiB, potentially due to the ultrasound measurements at the second timepoint being on a highly selected sample.

We show independent impacts of the fetal and maternal genomes on birthweight in two European (POPS, BiB-EUR) and one South Asian (BiB-SAS) cohorts. Fetal and maternal genetics together accounted for ∼11-12% of variance in birthweight in European ancestry groups, compared to ∼6% in the South Asian group.

In POPS, average PRS correlated with clinical groupings of observed birthweights, with larger babies having higher PRSs. Our findings suggest that PRS may be useful in discriminating between different phenotypes of small babies. The difference between observed and genetically-predicted birthweight was greater on average in babies designated as growth-restricted (IUGR) compared to those designated as constitutionally small (SGA).

### b. Results in the context of what is known

Current practices in dating pregnancies assume that all fetuses are equal in size at 11-14 weeks, when variation in fetal size is smallest. Thus all babies begin their fetal growth trajectory from a ‘standardized’ point, negating individual genetic influences on fetal size early in pregnancy^38,39^.

This may partly explain why PRSs explain more variance in EFW later in pregnancy in POPS, where GA was determined based on CRL in a first-trimester scan. In keeping with previous evidence^40^, we show a cumulative effect of fetal genetics that becomes more strongly associated with EFW as gestation progresses, despite reducing accuracy in scan measurements^41–44^. These observations may reflect the fact that EFW measured later in gestation is a better proxy for birthweight, though fetal PRS may become a pragmatic adjunct to predicting true fetal weight as birthweight GWASs become more powerful and it becomes possible to calculate fetal PRSs based on low-coverage NIPT data^45,46^.

In POPS, we show that while maternal PRS is not associated with EFW at mid-pregnancy, maternal genetic impacts accumulate faster than fetal genetic effects in the second half of pregnancy, although this finding was not replicated in BiB (see below). This may be attributable to the increasing importance of the maternal milieu in driving fetal growth, i.e., genetic propensity to insulin resistance^47,48^ and cardiovascular adaptations^49,50^.

Our results also suggest that polygenic risk for birthweight can be applied across ethnic groups. Maternal genetic effects on birthweight were comparable across cohorts, an important finding given the under-representation of Asian groups in genetic studies^51,52^. However, we note that some results in BiB-EUR and BiB-SAS differed qualitatively from each other and from POPS. For example, PRS_mother_ had a significant effect on EFW mid-pregnancy in BiB-SAS but not in BiB-EUR or POPS. Its effect size on EFW at week 22 was significantly bigger in BiB-SAS than BiB-EUR in the joint model (Table S7C; Δβ=0.088; p=0.026), whereas its effect size on birthweight did not differ significantly between ancestries (Table S9D; ; Δβ=0.138; p=0.17).

Replication in other cohorts and more powerful GWASs in South Asian will be required to confirm there are genuine differences in the impact of birthweight-associated genetic variation on EFW between ancestry groups. Another recent study reported associations of mid-pregnancy EFW with maternal as well as fetal birthweight genetic scores in BiB European and

South Asian women^53^; several factors could explain the differences in our findings, including how the PRS was constructed as well as differences in sample filtering.

### c. Clinical Implications

Our finding that genetic risk scores could be useful in differentiating IUGR from SGA is important because there is no gold standard for distinguishing IUGR from SGA^54^. Clinicians attempt to mitigate risks of stillbirth and neonatal morbidities associated with IUGR by recommending early delivery^55,56^, though this strategy can drive additional morbidity if applied to miscategorized SGA babies^57–59^. Conversely, mistaking IUGR for SGA can hinder surveillance and intervention that prevents stillbirth. Future studies should test whether PRSs derived from low-coverage NIPT data could improve diagnostic accuracy beyond existing clinical markers^37,60,61^.

### d. Research Implications

Future studies should consider how birthweight polygenic risk could be incorporated into clinical risk prediction tools for adverse perinatal outcomes. Our findings should also be replicated in cohorts with serial ultrasound data and in other ethnicities. Until new, cheaper approaches to non-invasive fetal sequencing are validated, using maternal data could mitigate fetal genetic testing constraints.

### e. Strengths and Limitations

We analyzed birthweight associations in two large, meticulously-phenotyped cohorts, POPS and BiB, with distinct ancestry groupings; POPS and BiB-EUR captured European ancestry groups and BiB-SAS captured South Asian ancestry groups. The availability of standardized, longitudinal whole-cohort EFW data from POPS alongside a comprehensive definition of IUGR is a major strength.

A limitation of research on ultrasonographic fetal weight estimates is that the error associated with measuring fetal size increases as pregnancy progresses^41,42^, such that fetal growth may be over-or under-estimated in late pregnancy^43^. Methods for deriving gestational age also differed between POPS and BiB. EFW measurements in BiB should be interpreted cautiously, as a limited, clinically-selected subset of BiB participants had ultrasound measurements for longitudinal analysis. Application of PRSs in BiB-SAS must account for the limited transferability of PRSs derived from GWAS of European ancestry participants to other ancestry groups^62^.

### f. Conclusions

Fetal and maternal genetic factors contribute independently to variation in fetal growth and birthweight, with their effects becoming more apparent as gestation progresses. Fetal genetic influences were detectable from mid-pregnancy, while maternal genetic effects appeared to strengthen later in gestation. We found that the discrepancy between observed and genetically-predicted birthweight may help distinguish growth-restricted from constitutionally-small infants. These findings provide new insight into genetic determinants of fetal growth and support further evaluation of polygenic approaches in perinatal risk assessment.

## Declarations

### Ethics approval and consent to participate

Informed consent was obtained from all POPS study participants. The study was approved by the Cambridgeshire 2 Research Ethics Committee (identification number 07/H0308/163). Ethics approval for BiB was obtained from the National Health Service Health Research Authority Yorkshire and the Humber (Bradford Leeds) Research Ethics Committee reference: 16/YH/0320 and 16/YH/0062). Mothers were consented to participate in the study upon attendance of a routine antenatal clinic appointment at the city’s primary maternity unit.

### Availability of data or materials

The raw data from the POPS study are not publicly available, due to the nature of the consent given by participants in the original study. Requests for POPS supplemental information, including raw data, can be made to Mrs Sheree Green-Molloy at the Department of Obstetrics and Gynaecology, Cambridge University, UK. Researchers can apply to access data from BiB (https://bib.vergelabs.dev/research/how-to-access-data/).

Data on birth weight has been contributed by the EGG Consortium using the UK Biobank Resource and has been downloaded from www.egg-consortium.org https://egg-consortium.org/birth-weight-2019.html

### CRediT Author Statement: Frances E Cates

Data curation, formal analysis, validation, writing - original draft, **Olivia Wootton:** Conceptualization, data curation, formal analysis, writing – original draft, **Jasmine X Kiley**: Methodology, validation, writing – review and editing, **Tabitha Wishlade:** Data curation, methodology, writing - review and editing, **Ulla Sovio:** Data curation, project administration, resources, writing – review and editing, **Gordon C S Smith:** Conceptualization, project administration, resources, writing – review and editing, **Jasmine A Mack:** Data curation, methodology, writing – review and editing, **D. Stephen Charnock-Jones:** Data curation, resources, writing – review and editing, **Hilary C Martin:** Conceptualization, methodology, project administration, supervision, writing – review and editing, **Catherine E Aiken:** Conceptualization, methodology, project administration, writing – review and editing.

## Data Availability

The raw data from the POPS study are not publicly available, due to the nature of the consent given by participants in the original study. Requests for POPS supplemental information, including raw data, can be made to Mrs Sheree Green-Molloy at the Department of Obstetrics and Gynaecology, Cambridge University, UK. Researchers can apply to access data from BiB (https://bib.vergelabs.dev/research/how-to-access-data/). Data on birth weight has been contributed by the EGG Consortium using the UK Biobank Resource and has been downloaded from www.egg-consortium.org https://egg-consortium.org/birth-weight-2019.html

## Acknowledgements

Born in Bradford is only possible because of the enthusiasm and commitment of the children and parents in BiB. We are grateful to all the participants, health professionals, schools and researchers who have made Born in Bradford happen.

## List of abbreviations

AGA: appropriate for gestational age
BiB-EUR: Born in Bradford participants of genetically-inferred European ancestry
BiB-SAS: Born in Bradford participants of genetically-inferred South Asian ancestry
D: deviation from genetically-predicted birthweight
EFW: estimated fetal weight
BW: birthweight
GA: gestational age
IUGR: intrauterine growth restriction
LGA: large for gestational age
PCs: genetic principal components
POPS: Pregnancy Outcomes Prediction Study
PRS: polygenic risk score
SGA: small for gestational age

