## Supplemental File 1 for "Contribution of maternal and fetal genetics to fetal weight throughout pregnancy and at birth"

Supplemental File 1: Supplementary Tables and Figures

**Supplementary Table 1**

|  | **POPS n** | **BiB (EUR) n** | **BiB (SAS) n** |
| --- | --- | --- | --- |
| BW_z_ Analysis | 2537 | 2075 | 2210 |
| EFW_z_ Analysis | **18-22 wks:** 2531  **26-30 wks:** 2515  **34-38 wks:** 2396 | 1358  **2 scans**: 728 | 1435  **2 scans**: 749 |

**Table S1. Descriptive statistics for birthweight and EFW analyses.** Descriptive table in which n represents the number of children in POPS, BiB (EUR), and BiB (SAS) groups included in birthweight and EFW analyses.

**Supplementary Table 2**

|  | **POPS n(%)** |  | **BiB-EUR n(%)** | **BiB-SAS n(%)** |
| --- | --- | --- | --- | --- |
| **Ethnicity** |  | **Ethnicity** |  |  |
| White | 2536 (100.0) | White | 2075 (100.0) |  |
| Missing | 1 (0) | Asian or Asian British |  | 2210 (100.0) |
| **IMD quartile** |  | **IMD quintile** |  |  |
| 1 (most deprived) | 635 (25.0) | 1 (most deprived) | 932 (44.9) | 1496 (67.7) |
| 2 | 627 (24.7) | 2 | 447 (21.5) | 304 (13.8) |
| 3 | 599 (23.5) | 3 | 377 (18.2) | 134 (6.1) |
| 4 (least deprived) | 561 (22.1) | 4 | 121 (5.8) | 11 (0.5) |
| Missing | 115 (4.5) | 5 (least deprived) | 73 (3.5) | 5 (0.2) |
| **Age** |  | **Age** |  |  |
| <20 | 88 (3.5) | <20 | 227 (10.9) | 82 (3.7) |
| 20-24.9 | 334 (13.2) | 20-24.9 | 549 (26.5) | 551 (24.9) |
| 25-29.9 | 793 (31.3) | 25-29.9 | 611 (29.4) | 759 (34.3) |
| 30-34.9 | 950 (37.4) | 30-34.9 | 444 (21.4) | 555 (25.1) |
| 35-39.9 | 330 (13) | 35-39.9 | 211 (10.2) | 234 (10.6) |
| ≥40 | 42 (1.7) | ≥40 | 33 (1.6) | 29 (1.3) |
| **BMI, kg/m^2^** |  | **BMI** |  |  |
| <25 | 1479 (58.3) | <25 | 856 (41.3) | 940 (42.5) |
| 25-29.9 | 708 (27.9) | 25-29.9 | 536 (25.8) | 542 (24.5) |
| 30-34.9 | 244 (9.6) | 30-34.9 | 266 (12.8) | 250 (11.3) |
| 35-39.9 | 73 (2.9) | 35-39.9 | 137 (6.6) | 84 (3.8) |
| ≥40 | 32 (1.3) | ≥40 | 67 (3.2) | 34 (1.5) |
| Missing | 1 (0.0) | Missing | 213 (10.3) | 360 (16.3) |
| **Smoking status** |  | **Smoking status** |  |  |
| Smoker during pregnancy | 116 (4.6) | Smoker during pregnancy | 600 (28.9) | 67 (3.0) |
| Missing | 0 (0) | Missing | 126 (3.2) | 267 (12.1) |
| **Alcohol consumption status** |  | **Alcohol** **consumption status** |  |  |
| Does not drink alcohol | 2401 (94.6) | No alcohol consumption in pregnancy | 449 (21.6) | 9 (0.4) |
| Does drink alcohol | 135 (5.3) | Alcohol consumption in pregnancy | 881 (42.5) | 11 (0.5) |
| Missing | 1 (0.04) | Missing | 745 (35.9) | 2190 (99.1) |
| **Employment status** |  | **Employment status** |  |  |
| Unemployed | 9 (0.35) | Unemployed | 603 (29.1) | 1460 (66.1) |
| Employed | 2523 (99.4) | Employed | 1347 (64.9) | 487 (22.0) |
| Missing | 5 (0.20) | Missing | 125 (6.02) | 263 (11.9) |

**Table S2. Characteristics of POPS and BiB cohorts.** Descriptive table reporting socio-demographic characteristics in two UK cohorts, POPS and BiB.

**Supplementary Table 3**

| **Clinical Marker of Growth Restriction** | **Threshold Indicating Placental Dysfunction** |
| --- | --- |
| Fetal biometric ratios |  |
| HC:AC | Top decile |
| AC:FL | Bottom decile |
| Fetal growth velocity |  |
| AC growth (z-score change) | Bottom decile |
| Doppler flow velocimetry |  |
| Uterine artery PI | Top decile |
| Umbilical artery PI | Top decile |
| Maternal serum biomarkers  PAPP-A  AFP  sFlt1:PlGF ratio | <0.4 MoM at 12 weeks GA  >2.0 at 20 weeks GA  >38 at 36 weeks GA |

**Table S3. Identification of Intrauterine Growth Restriction in POPS.** In POPS, Sovio et al. used five ultrasonic indices to detect growth restriction in POPS: head circumference (HC), abdominal circumference (AC), femur length (FL), uterine artery pulsatility index (PI), and umbilical artery PI^37^. All measures were quantified as gestational age adjusted z-scores. Uterine artery PI was characterized as the mean PI of the left and right uterine arteries at the 20-week scan measurement. Umbilical artery PI, HC:AC ratio, and AC:FL ratio were characterized by the last measurement before birth. Growth velocity was characterized by the difference in AC z-score between the last scan before birth and the 20-week scan. For each of the 5 main indices, deciles were generated with the gestational age adjusted z-scores. Placental dysfunction was defined as the highest deciles (top 10%) of HC:AC ratio, uterine Doppler, and umbilical Doppler and as the lowest deciles (bottom 10%) of AC:FL ratio and AC growth velocity. In addition to ultrasonic biomarkers, the following maternal serum biomarkers have been identified in POPS: pregnancy-associated plasma protein A (PAPP-A), alpha fetoprotein (AFP), and the ratio of soluble fms-like tyrosine kinase 1 (sFlt-1) to placental growth factor (PlGF). Growth restriction is indicated by PAPP-A<0.4 at 12 weeks GA^21^, AFP>2.0 at 20 weeks GA^21^, or sFlt:PlGF>38 at 36 weeks GA^22^. Having at least one ultrasonic biomarker, one maternal serum biomarker, and being designated as SGA (birthweight<10th centile) indicated fetal growth restriction in POPS.

**Supplementary Table 4A**

|  | **POPS** |  |  |  |  |  | **BiB (EUR)** | | | | | | **BiB (SAS)** | | | | | |
| --- | --- | --- | --- | --- | --- | --- | --- | --- | --- | --- | --- | --- | --- | --- | --- | --- | --- | --- |
| Coefficient | Estimate | SE | CI Lower | CI Upper | P-value | Q-value FDR | Estimate | SE | CI Lower | CI Upper | P-value | Q-value FDR | Estimate | SE | CI Lower | CI Upper | P-value | Q-value FDR |
| (Intercept) | -3.878 | 0.531 | -4.92 | -2.836 | 2.9e-13 |  | -0.048 | 0.15 | -0.343 | 0.246 | 0.747 |  | -0.689 | 0.19 | -1.061 | -0.317 | 2.81E-04 |  |
| PRS_fetus_ | 0.154 | 0.017 | 0.12 | 0.187 | 4.0e-19 | 1.2e-18 | 0.157 | 0.033 | 0.092 | 0.222 | 1.95e-06 | 2.84e-05 | 0.07 | 0.034 | 0.004 | 0.137 | 0.037 | 0.1056 |
| GA at scan | 0.003 | 0.001 | 0 | 0.005 | 0.034 | 0.034 | 0 | 0.003 | -0.006 | 0.006 | 0.94 | 0.9404 | -0.006 | 0.003 | -0.012 | -0.001 | 0.03 | 0.1056 |
| Maternal age | 0.008 | 0.003 | 0.002 | 0.014 | 0.009 |  | 0.021 | 0.005 | 0.01 | 0.031 | 7.13E-05 |  | 0.028 | 0.006 | 0.016 | 0.041 | 1.22E-05 |  |
| Maternal height | 0.017 | 0.002 | 0.012 | 0.022 | 1.7e-11 |  |  |  |  |  |  |  |  |  |  |  |  |  |
| Paternal height | 0.005 | 0.002 | 0 | 0.009 | 0.031 |  |  |  |  |  |  |  |  |  |  |  |  |  |
| PC1 | 3.058 | 1.177 | 0.752 | 5.365 | 0.009 |  | -10.289 | 7.184 | -24.37 | 3.792 | 0.152 |  | 3.647 | 3.283 | -2.788 | 10.081 | 0.267 |  |
| PC2 | 1.892 | 1.186 | -0.433 | 4.217 | 0.111 |  | -0.404 | 7.424 | -14.955 | 14.147 | 0.957 |  | -1.866 | 1.656 | -5.112 | 1.38 | 0.26 |  |
| PC3 | -2.028 | 1.168 | -4.318 | 0.262 | 0.083 |  | 1.178 | 5.432 | -9.469 | 11.824 | 0.828 |  | 0.874 | 1.636 | -2.332 | 4.08 | 0.593 |  |
| PC4 | 0.24 | 1.191 | -2.094 | 2.575 | 0.840 |  | -6.554 | 5.478 | -17.29 | 4.182 | 0.231 |  | 1.515 | 2.011 | -2.426 | 5.456 | 0.451 |  |
| PC5 | 0.413 | 1.17 | -1.881 | 2.707 | 0.724 |  | 1.454 | 4.345 | -7.061 | 9.97 | 0.738 |  | 1.597 | 2.378 | -3.064 | 6.257 | 0.502 |  |
| PC6 | -0.556 | 1.18 | -2.869 | 1.757 | 0.638 |  | 1.549 | 3.422 | -5.157 | 8.255 | 0.651 |  | 2.608 | 2.915 | -3.105 | 8.321 | 0.371 |  |
| PC7 | -0.246 | 1.17 | -2.54 | 2.047 | 0.833 |  | 0.964 | 3.044 | -5.003 | 6.93 | 0.752 |  | -1.994 | 1.967 | -5.848 | 1.861 | 0.311 |  |
| PC8 | -0.574 | 1.161 | -2.85 | 1.702 | 0.621 |  | 0.662 | 3.107 | -5.428 | 6.752 | 0.831 |  | -5.264 | 5.978 | -16.982 | 6.453 | 0.379 |  |
| PC9 | 0.967 | 1.166 | -1.318 | 3.252 | 0.407 |  | 0.969 | 2.916 | -4.746 | 6.685 | 0.74 |  | -4.098 | 2.779 | -9.544 | 1.348 | 0.14 |  |
| PC10 | -1.49 | 1.167 | -3.777 | 0.797 | 0.202 |  | 5.521 | 3.194 | -0.739 | 11.781 | 0.084 |  | 1.681 | 2.131 | -2.495 | 5.857 | 0.43 |  |
| PRS_fetus_:GA | 0.005 | 0.001 | 0.002 | 0.007 | 7.6e-05 | 1.1e-04 | 0.009 | 0.003 | 0.003 | 0.015 | 0.004 | 0.0127 | 0.004 | 0.003 | -0.001 | 0.01 | 0.131 | 0.2221 |

**Table S4A. Linear mixed effects model of longitudinal association between PRS_fetus_ and EFW_z_ with continuous GA.** Gestational age (GA) was modeled as a continuous variable to assess changes across gestation in the association of PRS_fetus_ with EFW. Estimates are presented with standard errors (SE), 95% confidence intervals (CIs), p-values, and q-values. False discovery rate correction was applied using the Benjamini-Hochberg method to generate q-values. **Model:** 2

**Supplementary Table 4B**

|  | **POPS** |  |  |  |  |  | **BiB (EUR)** | | | | | | **BiB (SAS)** | | | | | |
| --- | --- | --- | --- | --- | --- | --- | --- | --- | --- | --- | --- | --- | --- | --- | --- | --- | --- | --- |
| Coefficient | Estimate | SE | CI Lower | CI Upper | P-value | Q-value FDR | Estimate | SE | CI Lower | CI Upper | P-value | Q-value FDR | Estimate | SE | CI Lower | CI Upper | P-value | Q-value FDR |
| (Intercept) | -4.796 | 0.56 | -5.893 | -3.699 | 1.0e-17 |  | -0.071 | 0.154 | -0.372 | 0.23 | 0.642 |  | -0.678 | 0.19 | -1.051 | -0.305 | 3.64e-04 |  |
| PRS_mother_ | 0.032 | 0.018 | -0.004 | 0.068 | 0.081 | 0.121 | 0.046 | 0.034 | -0.02 | 0.112 | 1.71e-01 | 0.2903 | 0.058 | 0.035 | -0.01 | 0.127 | 0.095 | 0.1792 |
| GA at scan | 0.002 | 0.001 | -0.001 | 0.004 | 0.127 | 0.127 | 0.001 | 0.003 | -0.005 | 0.007 | 0.74 | 0.9146 | -0.006 | 0.003 | -0.012 | 0 | 0.034 | 0.1056 |
| Maternal age | 0.008 | 0.003 | 0.001 | 0.014 | 0.019 |  | 0.022 | 0.005 | 0.012 | 0.032 | 3.26E-05 |  | 0.028 | 0.006 | 0.015 | 0.04 | 1.91E-05 |  |
| Maternal height | 0.018 | 0.003 | 0.013 | 0.023 | 2.5e-11 |  |  |  |  |  |  |  |  |  |  |  |  |  |
| Paternal height | 0.009 | 0.002 | 0.004 | 0.014 | 1.5e-04 |  |  |  |  |  |  |  |  |  |  |  |  |  |
| PC1 | 2.97 | 1.258 | 0.504 | 5.436 | 0.018 |  | -10.709 | 7.339 | -25.092 | 3.675 | 0.145 |  | 4.488 | 3.263 | -1.907 | 10.883 | 0.169 |  |
| PC2 | 1.31 | 1.233 | -1.107 | 3.727 | 0.288 |  | -1.205 | 7.582 | -16.065 | 13.655 | 0.874 |  | -2.026 | 1.657 | -5.274 | 1.222 | 0.222 |  |
| PC3 | -3.202 | 1.296 | -5.742 | -0.662 | 0.013 |  | 1.3 | 5.553 | -9.583 | 12.183 | 0.815 |  | 0.642 | 1.639 | -2.571 | 3.855 | 0.695 |  |
| PC4 | 0.457 | 1.242 | -1.977 | 2.89 | 0.713 |  | -6.108 | 5.596 | -17.076 | 4.86 | 0.275 |  | 1.716 | 2.014 | -2.231 | 5.663 | 0.394 |  |
| PC5 | 0.364 | 1.219 | -2.025 | 2.754 | 0.765 |  | 0.4 | 4.434 | -8.29 | 9.091 | 0.928 |  | 1.608 | 2.383 | -3.063 | 6.28 | 0.5 |  |
| PC6 | -1.023 | 1.231 | -3.435 | 1.389 | 0.406 |  | 2.43 | 3.493 | -4.417 | 9.277 | 0.487 |  | 2.348 | 2.921 | -3.376 | 8.072 | 0.421 |  |
| PC7 | -0.979 | 1.221 | -3.372 | 1.415 | 0.423 |  | 0.032 | 3.104 | -6.053 | 6.117 | 0.992 |  | -1.721 | 1.968 | -5.578 | 2.136 | 0.382 |  |
| PC8 | -0.298 | 1.205 | -2.66 | 2.063 | 0.804 |  | 0.303 | 3.175 | -5.919 | 6.526 | 0.924 |  | -5.16 | 5.987 | -16.894 | 6.574 | 0.389 |  |
| PC9 | 0.674 | 1.212 | -1.701 | 3.048 | 0.578 |  | 0.326 | 2.976 | -5.508 | 6.159 | 0.913 |  | -3.989 | 2.789 | -9.455 | 1.477 | 0.153 |  |
| PC10 | -1.461 | 1.212 | -3.836 | 0.915 | 0.228 |  | 6.645 | 3.256 | 0.264 | 13.026 | 0.041 |  | 1.774 | 2.133 | -2.407 | 5.954 | 0.406 |  |
| PRS_mother_:GA | 0.009 | 0.001 | 0.006 | 0.011 | 1.1e-11 | 3.4e-11 | 0.01 | 0.003 | 0.004 | 0.016 | 9.39E-04 | 0.004 | 0.004 | 0.003 | -0.002 | 0.01 | 0.226 | 0.2754 |

**Table S4B. Linear mixed effects model of longitudinal association between PRS_mother_ and EFW_z_ with continuous GA.** Gestational age (GA) was modeled as a continuous variable to assess changes across gestation in the association of PRS_mother_ with EFW. Estimates are presented with standard errors (SE), 95% confidence intervals (CIs), and q-values. False discovery rate correction was applied using the Benjamini-Hochberg method. **Model:** 3

**Supplementary Table 4C**

|  | **POPS** |  |  |  |  |  | **BiB (EUR)** | | | | | | **BiB (SAS)** | | | | | |
| --- | --- | --- | --- | --- | --- | --- | --- | --- | --- | --- | --- | --- | --- | --- | --- | --- | --- | --- |
| Coefficient | Estimate | SE | CI Lower | CI Upper | P-value | Q-value FDR | Estimate | SE | CI Lower | CI Upper | P-value | Q-value FDR | Estimate | SE | CI Lower | CI Upper | P-value | Q-value FDR |
| (Intercept) | -3.981 | 0.555 | -5.07 | -2.892 | 7.7e-13 |  | -0.054 | 0.15 | -0.349 | 0.24 | 0.719 |  | -0.674 | 0.19 | -1.046 | -0.302 | 3.83E-04 |  |
| PRS_fetus_ | 0.156 | 0.019 | 0.119 | 0.192 | 4.2e-17 | 2.1e-16 | 0.155 | 0.034 | 0.089 | 0.222 | 5.01E-06 | 2.84E-05 | 0.06 | 0.035 | -0.009 | 0.128 | 0.089 | 0.1792 |
| PRS_mother_ | -0.002 | 0.019 | -0.038 | 0.034 | 0.915 | 0.915 | 0.008 | 0.034 | -0.058 | 0.075 | 0.808 | 0.9146 | 0.042 | 0.036 | -0.029 | 0.113 | 0.243 | 0.2754 |
| GA at scan | 0.002 | 0.001 | 0 | 0.004 | 0.110 | 0.137 | 0.001 | 0.003 | -0.005 | 0.006 | 0.861 | 0.9146 | -0.006 | 0.003 | -0.012 | -0.001 | 0.032 | 0.1056 |
| Maternal age | 0.008 | 0.003 | 0.001 | 0.014 | 0.020 |  | 0.021 | 0.005 | 0.011 | 0.031 | 6.07E-05 |  | 0.028 | 0.006 | 0.015 | 0.04 | 1.75E-05 |  |
| Maternal height | 0.016 | 0.003 | 0.011 | 0.021 | 5.6e-10 |  |  |  |  |  |  |  |  |  |  |  |  |  |
| Paternal height | 0.006 | 0.002 | 0.001 | 0.011 | 0.012 |  |  |  |  |  |  |  |  |  |  |  |  |  |
| PC1 | 2.682 | 1.23 | 0.272 | 5.093 | 0.029 |  | -10.139 | 7.183 | -24.217 | 3.939 | 0.158 |  | 3.628 | 3.278 | -2.797 | 10.052 | 0.268 |  |
| PC2 | 1.689 | 1.222 | -0.707 | 4.084 | 0.167 |  | -0.288 | 7.423 | -14.836 | 14.26 | 0.969 |  | -1.879 | 1.653 | -5.119 | 1.361 | 0.256 |  |
| PC3 | -2.52 | 1.277 | -5.023 | -0.017 | 0.048 |  | 1.023 | 5.434 | -9.627 | 11.674 | 0.851 |  | 0.749 | 1.635 | -2.456 | 3.954 | 0.647 |  |
| PC4 | 0.312 | 1.223 | -2.086 | 2.71 | 0.799 |  | -6.767 | 5.477 | -17.502 | 3.969 | 0.217 |  | 1.63 | 2.009 | -2.307 | 5.567 | 0.417 |  |
| PC5 | 0.052 | 1.199 | -2.297 | 2.401 | 0.965 |  | 1.422 | 4.343 | -7.091 | 9.935 | 0.743 |  | 1.755 | 2.377 | -2.903 | 6.414 | 0.46 |  |
| PC6 | -0.855 | 1.214 | -3.234 | 1.525 | 0.481 |  | 1.588 | 3.422 | -5.118 | 8.295 | 0.643 |  | 2.463 | 2.912 | -3.245 | 8.171 | 0.398 |  |
| PC7 | -0.482 | 1.206 | -2.846 | 1.882 | 0.689 |  | 0.997 | 3.043 | -4.967 | 6.962 | 0.743 |  | -1.916 | 1.964 | -5.766 | 1.933 | 0.329 |  |
| PC8 | -0.168 | 1.185 | -2.489 | 2.154 | 0.887 |  | 0.839 | 3.108 | -5.253 | 6.931 | 0.787 |  | -5.394 | 5.969 | -17.093 | 6.305 | 0.366 |  |
| PC9 | 0.524 | 1.194 | -1.816 | 2.864 | 0.661 |  | 0.92 | 2.915 | -4.793 | 6.634 | 0.752 |  | -3.834 | 2.781 | -9.285 | 1.616 | 0.168 |  |
| PC10 | -1.388 | 1.191 | -3.723 | 0.946 | 0.244 |  | 5.435 | 3.194 | -0.824 | 11.695 | 0.089 |  | 1.588 | 2.128 | -2.582 | 5.759 | 0.455 |  |
| PRS_fetus_:GA | 0.004 | 0.001 | 0.001 | 0.006 | 0.005 | 0.008 | 0.007 | 0.003 | 0.001 | 0.013 | 0.032 | 0.0596 | 0.004 | 0.003 | -0.002 | 0.01 | 0.225 | 0.2754 |
| PRS_mother_:GA | 0.008 | 0.001 | 0.005 | 0.011 | 6.5e-10 | 1.6e-09 | 0.008 | 0.003 | 0.002 | 0.014 | 0.007 | 0.0178 | 0.003 | 0.003 | -0.004 | 0.009 | 0.409 | 0.4095 |

**Table S4C. Joint model of longitudinal associations between PRS_fetus_, PRS_mother_, and EFW_z_ with continuous GA.** Gestational age (GA) was modeled as a continuous variable to assess changes across gestation in the associations of PRS_mother_ and PRS_fetus_ with EFW. Estimates are presented with standard errors (SE), 95% confidence intervals (CIs), p-values, and q-values. False discovery rate correction was applied using the Benjamini-Hochberg method to generate q-values. **Model:** 4

**Supplementary Table 4D**

|  | **POPS** |  |  |  |  |  |
| --- | --- | --- | --- | --- | --- | --- |
| Coefficients | Beta | SE | 95% CI Lower | 95% CI Upper | P-value | Q-value FDR |
| (Intercept) | -3.535 | 0.556 | -4.624 | -2.446 | 2.0e-10 |  |
| PRS_fetus_ | 0.148 | 0.019 | 0.112 | 0.185 | 1.3e-15 | 6.5e-15 |
| PRS_mother_ | -0.003 | 0.019 | -0.039 | 0.034 | 0.885 | 0.885 |
| GA | 0.002 | 0.001 | 0 | 0.005 | 0.090 | 0.112 |
| Maternal age at booking | 0.007 | 0.004 | 0 | 0.014 | 0.045 |  |
| Maternal height | 0.008 | 0.003 | 0.003 | 0.014 | 0.002 |  |
| Paternal height | 0.006 | 0.002 | 0.002 | 0.011 | 0.006 |  |
| Maternal age at termination of education | 0.002 | 0.005 | -0.008 | 0.012 | 0.734 |  |
| Maternal weight at first scan | 0.011 | 0.001 | 0.009 | 0.014 | 2.6e-18 |  |
| Low socioeconomic status score | -0.069 | 0.023 | -0.114 | -0.024 | 0.003 |  |
| PC1 | 1.93 | 1.282 | -0.583 | 4.443 | 0.132 |  |
| PC2 | 1.451 | 1.229 | -0.958 | 3.86 | 0.238 |  |
| PC3 | -1.904 | 1.288 | -4.428 | 0.621 | 0.139 |  |
| PC4 | -0.041 | 1.227 | -2.445 | 2.363 | 0.973 |  |
| PC5 | 0.297 | 1.2 | -2.054 | 2.648 | 0.805 |  |
| PC6 | -0.911 | 1.222 | -3.306 | 1.484 | 0.456 |  |
| PC7 | -0.35 | 1.208 | -2.717 | 2.018 | 0.772 |  |
| PC8 | -0.697 | 1.184 | -3.017 | 1.624 | 0.556 |  |
| PC9 | 0.425 | 1.193 | -1.914 | 2.763 | 0.722 |  |
| PC10 | -1.226 | 1.191 | -3.56 | 1.108 | 0.303 |  |
| PRS_fetus_:GA | 0.004 | 0.001 | 0.001 | 0.006 | 0.004 | 0.007 |
| PRS_mother_GA | 0.008 | 0.001 | 0.005 | 0.01 | 2.4e-09 | 5.9e-09 |

**Table S4D. Fully-adjusted fixed-effects estimates from joint PRS model on EFW_z_ across gestation in POPS.** Gestational age (GA) was modeled as a continuous variable to assess changes across gestation in the associations of PRS_mother_ and PRS_fetus_ with EFW. Estimates are presented with standard errors (SE), 95% confidence intervals (CIs), p-values, and q-values. False discovery rate (FDR) correction was applied using the Benjamini-Hochberg method to generate q-values. **Adjustments:** maternal age at booking, maternal height, paternal height, maternal age at termination of education, maternal weight at first scan, low socioeconomic score. **Model:** 4

**Supplementary Table 5**

| **Cohort** | **Contrast** | **Estimate** | **Z** | **P-value** |
| --- | --- | --- | --- | --- |
| POPS | β_fetus:GA_– β_mother:GA_ | -0.004 | -2.152 | 0.031 |
| BiB-EUR | β_fetus:GA_-β_mother:GA_ | -0.0016 | -0.330 | 0.7418 |
| BiB-SAS | β_fetus:GA_-β_mother:GA_ | 0.0011 | 0.228 | 0.8198 |

**Table S5. Wald test of difference in maternal vs. fetal PRS effect interaction with continuous GA.** Differences in the longitudinal change of maternal vs. fetal PRS associations with EFW were evaluated using a linear contrast of the PRS x GA interaction terms from the linear-mixed effects model with continuous GA. Specifically, the contrast tested β_fetus:GA_– β_mother:GA_, which represents the rate of change in PRS effects across gestation between fetal and maternal PRS. Standard errors for the contrast were derived using the delta method based on the fixed effect variance-covariance matrix, and statistical inference was performed using a Wald z-test. **Model:** 4

**Supplementary Figure 1**

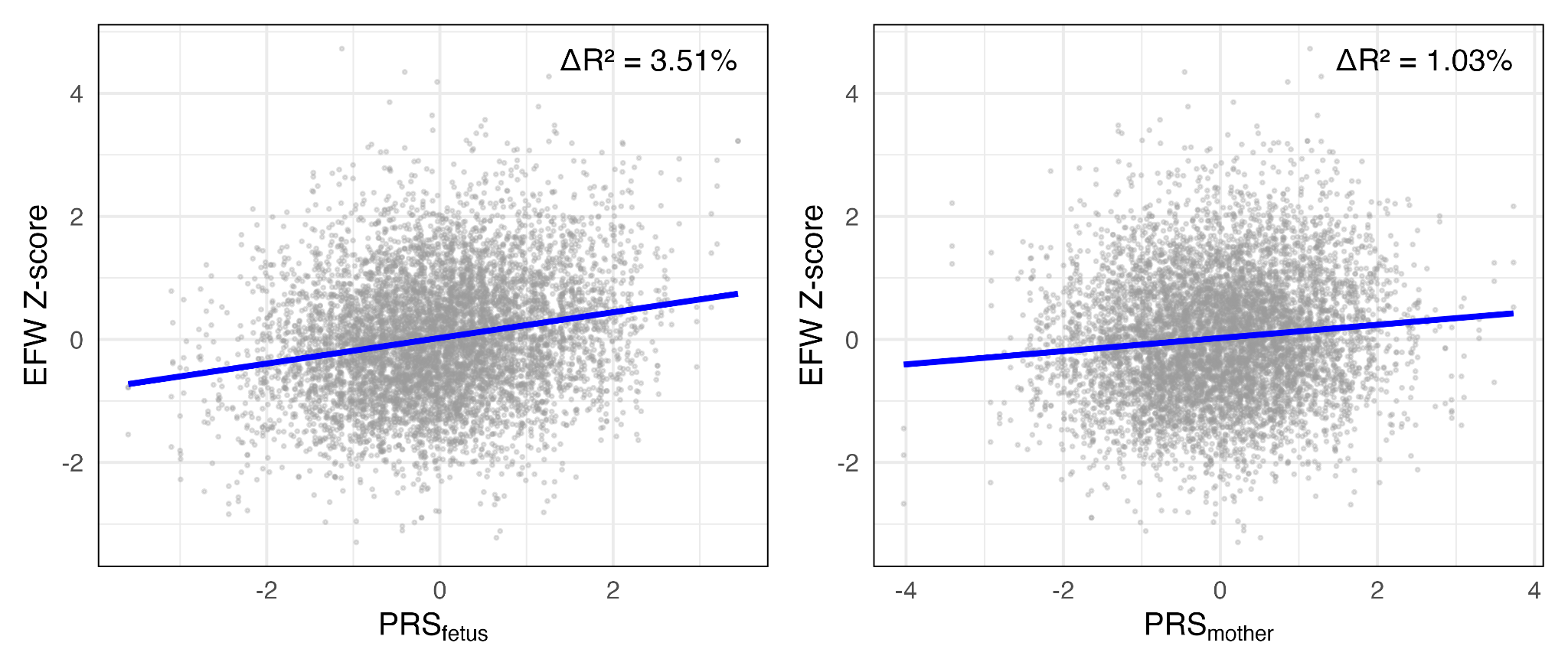

**Associations of fetal and maternal polygenic scores with estimated fetal weight in a linear mixed-effects model with continuous gestational age (POPS).** This plot shows associations between polygenic scores (PRS) and standardized EFW in a joint linear mixed-effects model with continuous gestational age in POPS using Model 4. The left panel shows the association with the fetal PRS (PRS_fetus_), and the right panel shows the association with the maternal PRS (PRS_mother_). Points represent individual observations and the blue line indicates the fitted linear regression. ΔR² represents the incremental variance in EFW explained by each PRS beyond covariates, calculated as the difference between the marginal R² of the full model and the covariates-only model.

**Supplementary Table 6A**

|  | **POPS** |  |  |  |
| --- | --- | --- | --- | --- |
| Model | Marginal R^2^ | Conditional R^2^ | ΔR^2^ | AIC |
| Model 1: Covariates only | 2.97% | 52.93% | NA | 18682.25 |
| Model 2: Fetal PRS | 6.48% | 53.09% | 3.51% | 18556.34 |
| Model 3: Maternal PRS | 4.01% | 53.41% | 1.03% | 18628.54 |
| Model 4: Maternal + Fetal PRS | 6.93% | 53.47% | 3.95% | 18532.41 |
| Model 5: Maternal$\times$ Fetal PRS | 6.98% | 53.48% | 4.01% | 18538.58 |

**Table S6A. Comparison of model performance for predicting EFW_z_ in POPS with continuous GA.** In POPS, we generated R^2^ and AIC values to represent the variance in EFW explained by the covariate-only (Model 1), fetal (Model 2), maternal (Model 3), joint (Model 4), and interactive (Model 5) longitudinal models with continuous GA. All ΔR^2^ values were calculated in reference to the covariate-only model. Marginal R^2^ values represented the variance in EFW_z_ explained solely by fixed effects, while conditional R^2^ values represented the variance explained by both fixed and random effects. **Adjustments:** maternal height, paternal height, maternal age, PCs 1-10. **Models:** 1-5

**Supplementary Table 6B**

|  | **POPS** |
| --- | --- |
| Genetic Component | Variance explained % |
| Independent fetal effect (ΔR^2^ fetal PRS) | 2.92 |
| Independent maternal effect (ΔR^2^ maternal PRS) | 0.45 |

**Table S6B. Variance decomposition for EFW_z_ in POPS with continuous GA.** We calculated the proportion of variance in EFW_z_ explained independently by PRS_mother_ and PRS_fetus_. Independent fetal and maternal effects were calculated by subtracting the variance explained by the fetal (Model 2) and maternal (Model 3) models from the joint (Model 4) model. **Adjustments:** maternal height, paternal height, maternal age at booking, PCs 1-10. **Models:** 2-4

**Supplementary Table 6C**

| **Comparison** | **Chi sq** | **df** | **P-value** |
| --- | --- | --- | --- |
| Joint (Model 9) vs interactive (Model 10) | 2.30 | 1 | 0.129 |

**Table S6C. Likelihood ratio test comparing joint and interactive linear mixed effects models for predicting EFW_z_ with continuous GA.** We conducted a likelihood ratio test to evaluate whether the interaction of PRS_mother_ and PRS_fetus_ improved model fit for predicting EFW_z_, comparing the joint (Model 4) and interactive (Model 5) models. **Adjustments:** maternal age at booking, maternal height, paternal height, PCs 1-10. **Models:** 4-5

**Supplementary Table 7A**

|  | **BiB (EUR)** | | | | | | **BiB (SAS)** | | | | | |
| --- | --- | --- | --- | --- | --- | --- | --- | --- | --- | --- | --- | --- |
| Coefficient | Beta | SE | CI Lower | CI Upper | P-value | Q-value FDR | Beta | SE | CI Lower | CI Upper | P-value | Q-value FDR |
| (Intercept) | 0.163 | 0.131 | -0.093 | 0.419 | 0.212 |  | -0.211 | 0.158 | -0.521 | 0.1 | 0.184 |  |
| PRS_fetus_ | 0.114 | 0.027 | 0.06 | 0.167 | 3.11e-05 | 8.83e-05 | 0.066 | 0.027 | 0.014 | 0.119 | 0.013 | 0.024 |
| Maternal age at booking | 0.011 | 0.005 | 0.002 | 0.02 | 0.017 |  | 0.013 | 0.006 | 0.002 | 0.024 | 0.019 |  |
| PC1 | -3.399 | 6.293 | -15.743 | 8.591 | 0.545 |  | 2.048 | 2.808 | -3.459 | 7.556 | 0.466 |  |
| PC2 | 4.237 | 6.26 | -8.044 | 16.518 | 0.499 |  | -2.065 | 1.436 | -4.882 | 0.752 | 0.151 |  |
| PC3 | 5.27 | 4.482 | -3.523 | 14.063 | 0.24 |  | 0.284 | 1.42 | -2.502 | 3.07 | 0.841 |  |
| PC4 | -8.484 | 4.905 | -18.106 | 1.138 | 0.084 |  | 2.345 | 1.74 | -1.068 | 5.758 | 0.178 |  |
| PC5 | -2.245 | 3.793 | -9.686 | 5.197 | 0.554 |  | -0.886 | 2.179 | -5.161 | 3.389 | 0.684 |  |
| PC6 | 1.646 | 3.029 | -4.296 | 7.587 | 0.587 |  | 3.866 | 2.494 | -1.027 | 8.759 | 0.121 |  |
| PC7 | 0.689 | 2.821 | -4.846 | 6.223 | 0.807 |  | -1.596 | 1.71 | -4.95 | 1.759 | 0.351 |  |
| PC8 | -1.62 | 2.592 | -6.704 | 3.465 | 0.532 |  | -0.715 | 4.851 | -10.231 | 8.8 | 0.883 |  |
| PC9 | 0.125 | 2.551 | -4.879 | 5.13 | 0.961 |  | -3.889 | 2.435 | -8.666 | 0.889 | 0.111 |  |
| PC10 | 2.661 | 2.701 | -2.639 | 7.96 | 0.325 |  | 1.167 | 2.018 | -2.791 | 5.125 | 0.563 |  |

**Table S7A. Cross-sectional associations between PRS_fetus_ and EFW at 18-22 weeks gestation in Born in Bradford (BiB).** We constructed a linear model regressing EFW_z_ across PRS_fetus_ (Model 7) to assess the association of EFW_z_ with fetal polygenic risk at 18-22 weeks gestation. Models were adjusted for maternal age at booking and the first 10 genetic principal components (PCs). Results are presented as beta coefficients, standard errors (SE), 95% confidence intervals (CI), nominal P-values, and false discovery rate (FDR)-adjusted q-values. **Model:** 7

**Supplementary Figures 2A&B**

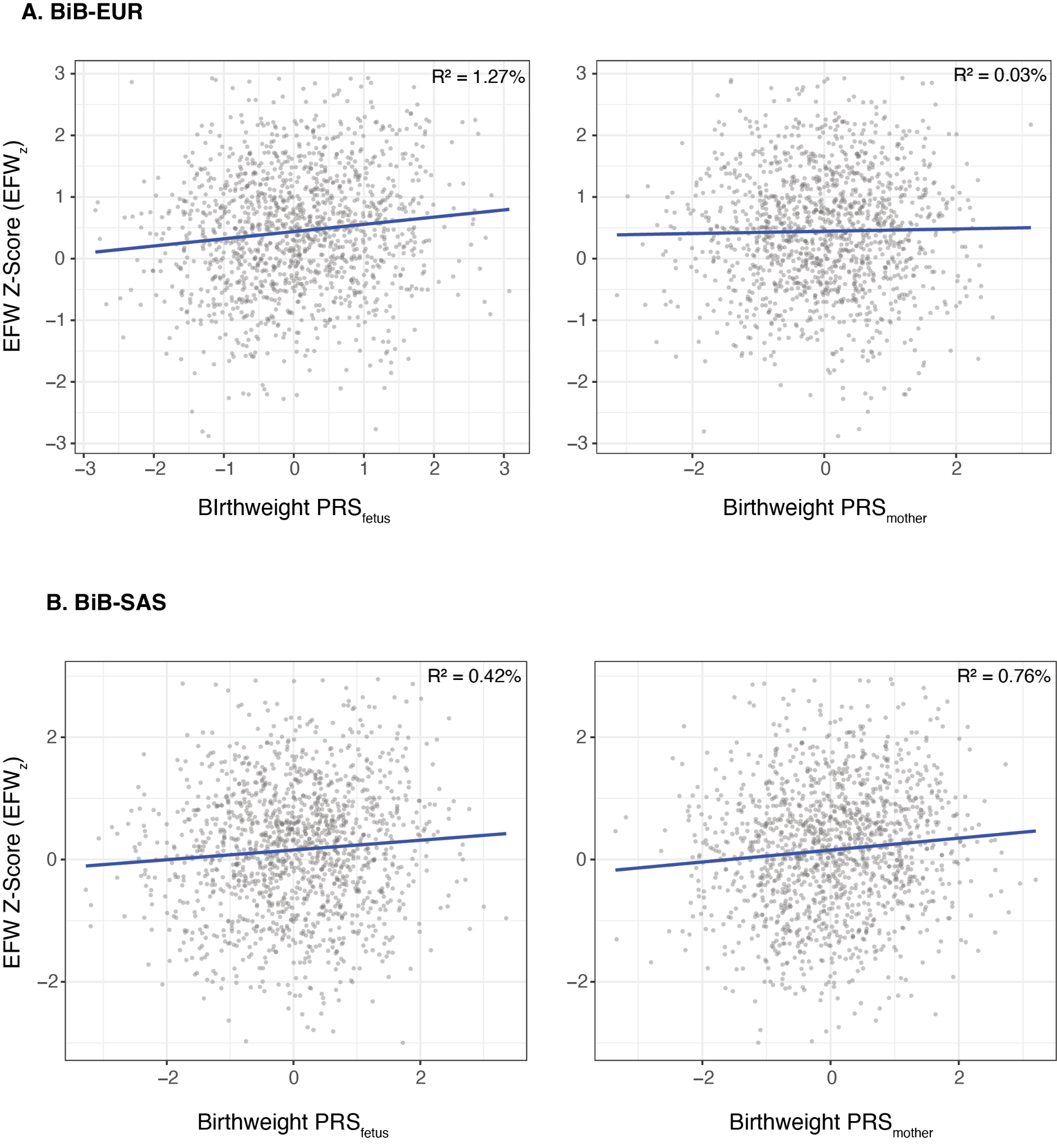

**Figures S2A&B. Cross-sectional associations between PRS and EFW_z_ at 18-22 weeks gestation in Born in Bradford (BiB)**. Each panel shows a linear regression of standardised EFW on a birthweight PRS, the left panel shows association with fetal PRS for own birthweight (PRS_fetus_) and the right panel shows association with the mother’s PRS for offspring birthweight (PRS_mother_) (models 7-8, Methods). The blue line indicates the fitted regression, R^2^ is the incremental variance explained in EFW by each PRS over and above covariates. EUR: genetically-inferred European ancestry (N=1,358); BiB-SAS: genetically-inferred South Asian ancestry (N=1,435)

**Supplementary Table 7B**

|  | **BiB (EUR)** | | | | | | **BiB (SAS)** | | | | | |
| --- | --- | --- | --- | --- | --- | --- | --- | --- | --- | --- | --- | --- |
| Coefficient | Beta | SE | CI Lower | CI Upper | P-value | Q-value FDR | Beta | SE | CI Lower | CI Upper | P-value | Q-value FDR |
| (Intercept) | 0.154 | 0.131 | -0.104 | 0.412 | 0.242 |  | -0.2 | 0.158 | -0.51 | 0.11 | 0.207 |  |
| PRS_mother_ | 0.019 | 0.027 | -0.035 | 0.073 | 0.491 | 0.688 | 0.091 | 0.027 | 0.037 | 0.144 | 9.01e-0.4 | 0.006 |
| Maternal age at booking | 0.012 | 0.005 | 0.003 | 0.021 | 0.012 |  | 0.012 | 0.005 | 0.002 | 0.023 | 0.024 |  |
| PC1 | -3.381 | 6.332 | -16.253 | 8.946 | 0.589 |  | 2.367 | 2.791 | -3.107 | 7.841 | 0.396 |  |
| PC2 | 3.8 | 6.3 | -8.56 | 16.61 | 0.547 |  | -2.313 | 1.432 | -5.122 | 0.497 | 0.107 |  |
| PC3 | 5.298 | 4.513 | -3.554 | 14.151 | 0.241 |  | 0.339 | 1.418 | -2.442 | 3.121 | 0.811 |  |
| PC4 | -8.706 | 4.936 | -18.389 | 0.976 | 0.078 |  | 2.332 | 1.737 | -1.076 | 5.739 | 0.18 |  |
| PC5 | -2.775 | 3.815 | -10.259 | 4.71 | 0.467 |  | -0.835 | 2.176 | -5.103 | 3.433 | 0.701 |  |
| PC6 | 1.671 | 3.048 | -4.308 | 7.649 | 0.584 |  | 3.99 | 2.49 | -0.895 | 8.875 | 0.109 |  |
| PC7 | 0.223 | 2.837 | -5.342 | 5.788 | 0.937 |  | -1.597 | 1.706 | -4.944 | 1.75 | 0.35 |  |
| PC8 | -1.491 | 2.609 | -6.608 | 3.627 | 0.568 |  | -0.425 | 4.842 | -9.924 | 9.074 | 0.93 |  |
| PC9 | -0.111 | 2.567 | -5.148 | 4.925 | 0.965 |  | -3.753 | 2.432 | -8.524 | 1.017 | 0.123 |  |
| PC10 | 2.744 | 2.718 | -2.589 | 8.077 | 0.313 |  | 0.973 | 2.016 | -2.982 | 4.928 | 0.63 |  |

**Table S7B. Cross-sectional associations between PRS_mother_ and EFW at 18-22 weeks gestation in Born in Bradford (BiB).** We constructed a linear model regressing EFW_z_ across PRS_mother_ (Model 8) to assess the association of EFW_z_ with maternal polygenic risk at 18-22 weeks gestation. Models were adjusted for maternal age at booking and the first 10 genetic principal components (PCs). Results are presented as beta coefficients, standard errors (SE), 95% confidence intervals (CI), nominal P-values, and false discovery rate (FDR)-adjusted q-values. **Model:** 8

**Supplementary Table 7C**

|  | **BiB (EUR)** | | | | | | **BiB (SAS)** | | | | | |
| --- | --- | --- | --- | --- | --- | --- | --- | --- | --- | --- | --- | --- |
| Coefficient | Beta | SE | CI Lower | CI Upper | P-value | Q-value FDR | Beta | SE | CI Lower | CI Upper | P-value | Q-value FDR |
| (Intercept) | 0.164 | 0.131 | -0.092 | 0.421 | 0.209 |  | -0.203 | 0.158 | -0.513 | 0.107 | 0.2 |  |
| PRS_fetus_ | 0.116 | 0.028 | 0.061 | 0.171 | 3.79e-05 | 8.83e-05 | 0.045 | 0.028 | -0.01 | 0.1 | 0.107 | 0.131 |
| PRS_mother_ | -0.01 | 0.028 | -0.065 | 0.046 | 0.735 | 0.736 | 0.078 | 0.028 | 0.022 | 0.133 | 0.006 | 0.015 |
| Maternal age at booking | 0.011 | 0.005 | 0.002 | 0.02 | 0.017 |  | 0.013 | 0.005 | 0.002 | 0.023 | 0.022 |  |
| PC1 | -3.431 | 6.295 | -15.78 | 8.919 | 0.586 |  | 1.939 | 2.802 | -3.557 | 7.435 | 0.489 |  |
| PC2 | 4.288 | 6.264 | -8 | 16.577 | 0.494 |  | -2.192 | 1.434 | -5.004 | 0.621 | 0.127 |  |
| PC3 | 5.316 | 4.486 | -3.484 | 14.116 | 0.236 |  | 0.336 | 1.417 | -2.444 | 3.116 | 0.813 |  |
| PC4 | -8.463 | 4.907 | -18.089 | 1.162 | 0.085 |  | 2.31 | 1.736 | -1.096 | 5.715 | 0.184 |  |
| PC5 | -2.257 | 3.795 | -9.701 | 5.188 | 0.552 |  | -0.798 | 2.175 | -5.063 | 3.468 | 0.714 |  |
| PC6 | 1.652 | 3.03 | -4.292 | 7.595 | 0.586 |  | 3.961 | 2.489 | -0.921 | 8.844 | 0.112 |  |
| PC7 | 0.687 | 2.822 | -4.85 | 6.223 | 0.808 |  | -1.7 | 1.707 | -5.047 | 1.648 | 0.319 |  |
| PC8 | -1.635 | 2.593 | -6.723 | 3.452 | 0.528 |  | -0.537 | 4.84 | -10.032 | 8.957 | 0.912 |  |
| PC9 | 0.145 | 2.553 | -4.862 | 5.153 | 0.955 |  | -3.717 | 2.431 | -8.486 | 1.051 | 0.126 |  |
| PC10 | 2.655 | 2.702 | -2.646 | 7.957 | 0.326 |  | 0.914 | 0.914 | -3.039 | 4.867 | 0.65 |  |

**Table S7C. Cross-sectional associations from joint model regressing EFW (18-22 weeks gestation) on PRS_fetus_ and PRS_mother_ (BiB).** We constructed a joint linear model regressing EFW_z_ across the PRS_mother_ and PRS_fetus_ (Model 9) to assess the association of EFW_z_ with fetal and maternal polygenic risk at 18-22 weeks gestation. Models were adjusted for maternal age at booking and the first 10 genetic principal components (PCs). Results are presented as beta coefficients, standard errors (SE), 95% confidence intervals (CI), nominal P-values, and false discovery rate (FDR)-adjusted q-values. **Model:** 9

**Supplementary Table 8A**

|  | **BiB (EUR)** |  |  | **BiB (SAS)** |  |  |
| --- | --- | --- | --- | --- | --- | --- |
| Model | R^2^ | ΔR^2^ | AIC | R^2^ | ΔR^2^ | AIC |
| Model 6: Covariates only | 0.97% |  | 3826.8 | 1.74% |  | 4190.8 |
| Model 7: Fetal PRS | 2.24% | 1.27% | 3811.3 | 2.16% | 0.42% | 4186.6 |
| Model 8: Maternal PRS | 1% | 0.03% | 3828.4 | 2.5% | 0.76% | 4181.7 |
| Model 9: Maternal + Fetal PRS | 2.25% | 1.28% | 3813.2 | 2.68% | 0.94% | 4181.1 |
| Model 10: Maternal$\times$Fetal PRS | 2.29% | 1.32% | 3814.6 | 2.71% | 0.96% | 4182.7 |

**Table 7A: Comparison of model performance for predicting EFW_z_ in BiB.** In BiB, we generated R^2^ and AIC values to represent the variance in EFW explained by the covariate-only (Model 6), fetal (Model 7), maternal (Model 8), joint (Model 9), and interactive (Model 10) cross-sectional models (18-22 week EFW). All ΔR^2^ values were calculated in reference to the covariate-only model. **Adjustments:** maternal age, PCs 1-10. **Models:** 6-10

**Supplementary Table 8B**

|  | **BiB (EUR)** | **BiB (SAS)** |
| --- | --- | --- |
| Genetic Component | Variance explained % | Variance explained % |
| Independent fetal effect (ΔR^2^ fetal PRS) | 1.24 | 0.18 |
| Independent maternal effect (ΔR^2^ maternal PRS) | 0.01 | 0.52 |

**Table 8B: Variance decomposition for EFW_z_ in BiB.** In BiB, we calculated the proportion of variance in EFW_z_ explained independently by PRS_mother_ and PRS_fetus_ in the cross-sectional models (18-22 week EFW). Independent fetal and maternal effects were calculated by subtracting the variance explained by the fetal (Model 7) and maternal (Model 8) linear models from the joint (Model 9) model. **Adjustments:** maternal age at booking, PCs 1-10. **Models:** 7-9

**Supplementary Table 9A**

|  | **POPS** | | | | | | **BiB (EUR)** | | | | | | **BiB (SAS)** | | | | | |
| --- | --- | --- | --- | --- | --- | --- | --- | --- | --- | --- | --- | --- | --- | --- | --- | --- | --- | --- |
| Coefficient | Beta | SE | CI Lower | CI Upper | P-value | Q-value FDR | Beta | SE | CI Lower | CI Upper | P-value | Q-value FDR | Beta | SE | CI Lower | CI Upper | P-value | Q-value FDR |
| (Intercept) | -4.428 | 0.549 | -5.506 | -3.351 | 1.17e-15 |  | -0.702 | 0.0962 | -0.891 | -0.514 | 3.68e-13 |  | -1.157 | 0.108 | -1.368 | -0.945 | 3.77e-26 |  |
| PRS_fetus_ | 0.27 | 0.016 | 0.238 | 0.302 | 4.88e-59 | 3.41e-58 | 0.312 | 0.02 | 0.273 | 0.351 | 6.54e-53 | 4.58e-52 | 0.218 | 0.019 | 0.18 | 0.256 | 9.31e-29 | 6.52e-28 |
| Maternal height | 0.019 | 0.003 | 0.014 | 0.025 | 5.61e-14 |  |  |  |  |  |  |  |  |  |  |  |  |  |
| Paternal height | 0.007 | 0.002 | 0.003 | 0.012 | 0.002 |  |  |  |  |  |  |  |  |  |  |  |  |  |
| Maternal age at booking | -0.007 | 0.003 | -0.013 | 0 | 0.043 |  | 0.026 | 0.003 | 0.02 | 0.033 | 1.26e-14 |  | 0.025 | 0.004 | 0.017 | 0.032 | 7.01e-11 |  |
| PC1 | 0.522 | 1.218 | -1.866 | 2.91 | 0.668 |  | -8.095 | 4.516 | -16.951 | 0.76 | 0.073 |  | 1.612 | 1.949 | -2.211 | 5.434 | 0.408 |  |
| PC2 | -1.158 | 1.213 | -3.538 | 1.221 | 0.340 |  | -1.925 | 4.63 | -11.005 | 7.155 | 0.678 |  | -3.773 | 1.008 | -5.749 | -1.796 | 1.87e-04 |  |
| PC3 | -0.674 | 1.266 | -3.156 | 1.807 | 0.594 |  | 0.145 | 3.254 | -6.237 | 6.527 | 0.964 |  | -1.255 | 1.016 | -3.248 | 0.738 | 0.217 |  |
| PC4 | 0.599 | 1.216 | -1.785 | 2.984 | 0.622 |  | -3.045 | 3.625 | -10.155 | 4.065 | 0.401 |  | 0.646 | 1.201 | -1.708 | 3.001 | 0.59 |  |
| PC5 | 0.992 | 1.19 | -1.341 | 3.324 | 0.405 |  | 2.46 | 2.758 | -2.949 | 7.869 | 0.373 |  | -0.329 | 1.448 | -3.169 | 2.511 | 0.82 |  |
| PC6 | 0.817 | 1.204 | -1.544 | 3.178 | 0.497 |  | 0.274 | 2.257 | -4.153 | 4.7 | 0.904 |  | 1.724 | 1.74 | -1.688 | 5.136 | 0.322 |  |
| PC7 | -0.378 | 1.197 | -2.725 | 1.97 | 0.752 |  | 1.37 | 2.055 | -2.66 | 5.399 | 0.505 |  | -1.841 | 1.236 | -4.266 | 0.584 | 0.137 |  |
| PC8 | -1.091 | 1.175 | -3.395 | 1.213 | 0.353 |  | 0.042 | 1.922 | -3.728 | 3.812 | 0.983 |  | -7.025 | 3.393 | -13.678 | -0.372 | 0.039 |  |
| PC9 | -0.456 | 1.184 | -2.778 | 1.865 | 0.700 |  | 2.817 | 1.881 | -0.872 | 6.507 | 0.134 |  | -3.322 | 1.706 | -6.667 | 0.023 | 0.052 |  |
| PC10 | -2.895 | 1.181 | -5.211 | -0.58 | 0.014 |  | 4.899 | 2.001 | 0.974 | 8.824 | 0.014 |  | -2.854 | 1.446 | -5.691 | -0.017 | 0.049 |  |
| **Table S9A. Association of BW_z_ with PRS_fetus_ across cohorts.** Linear regression analyses examined the association between observed birthweight and fetal polygenic risk (PRS_fetus_) using a univariable model (Model 12). Results are presented as coefficients, beta values, standard errors (SE), 95% confidence intervals (CI), p-values, and q-values. Multiple testing was corrected using the Benjamini-Hochberg false discovery rate (FDR) method to generate q-values. POPS models were adjusted for maternal age at booking, maternal height, paternal height, and genetic principal components (PCs) 1-10, while BiB models were adjusted for maternal age at booking and PCs 1-10. **Model:** 12 | | | | | | | | | | | | | | | | | | |

**Supplementary Table 9B**

|  | **POPS** | | | | | | **BiB (EUR)** | | | | | | **BiB (SAS)** | | | | | |
| --- | --- | --- | --- | --- | --- | --- | --- | --- | --- | --- | --- | --- | --- | --- | --- | --- | --- | --- |
| Coefficient | Beta | SE | CI Lower | CI Upper | P-value | Q-value FDR | Beta | SE | CI Lower | CI Upper | P-value | Q-value FDR | Beta | SE | CI Lower | CI Upper | P-value | Q-value FDR |
| (Intercept) | -5.247 | 0.565 | -6.355 | -4.139 | 3.46e-20 |  | -0.758 | 0.099 | -0.952 | -0.564 | 3.05e-14 |  | -1.139 | 0.11 | -1.354 | -0.925 | 9.24e-25 |  |
| PRS_mother_ | 0.165 | 0.017 | 0.132 | 0.198 | 3.79e-22 | 6.63e-22 | 0.21 | 0.021 | 0.169 | 0.25 | 6.89e-24 | 1.21e-23 | 0.15 | 0.02 | 0.112 | 0.189 | 2.33e-14 | 4.07e-14 |
| Maternal height | 0.019 | 0.003 | 0.014 | 0.025 | 9.97e-13 |  |  |  |  |  |  |  |  |  |  |  |  |  |
| Paternal height | 0.012 | 0.002 | 0.007 | 0.017 | 8.96e-07 |  |  |  |  |  |  |  |  |  |  |  |  |  |
| Maternal age at booking | -0.007 | 0.003 | -0.014 | 0 | 0.042 |  | 0.028 | 0.003 | 0.021 | 0.035 | 1.06e-15 |  | 0.024 | 0.004 | 0.016 | 0.031 | 4.61e-10 |  |
| PC1 | 0.563 | 1.262 | -1.91 | 3.037 | 0.655 |  | -6.575 | 4.664 | -15.722 | 2.571 | 0.159 |  | 2.787 | 1.973 | -1.082 | 6.657 | 0.158 |  |
| PC2 | -1.586 | 1.256 | -4.049 | 0.876 | 0.207 |  | -3.715 | 4.781 | -13.092 | 5.661 | 0.437 |  | -4.425 | 1.024 | -6.433 | -2.417 | 1.62e-0.5 |  |
| PC3 | -1.053 | 1.311 | -3.623 | 1.518 | 0.422 |  | -1.264 | 3.362 | -7.857 | 5.33 | 0.707 |  | -1.269 | 1.032 | -3.292 | 0.754 | 0.219 |  |
| PC4 | 0.31 | 1.259 | -2.158 | 2.777 | 0.806 |  | -1.474 | 3.742 | -8.813 | 5.865 | 0.694 |  | 0.483 | 1.218 | -1.907 | 2.872 | 0.692 |  |
| PC5 | 0.857 | 1.233 | -1.56 | 3.275 | 0.487 |  | 1.681 | 2.848 | -3.904 | 7.265 | 0.555 |  | -0.51 | 1.47 | -3.393 | 2.372 | 0.728 |  |
| PC6 | 0.336 | 1.247 | -2.11 | 2.781 | 0.788 |  | 0.254 | 2.332 | -4.319 | 4.827 | 0.913 |  | 2.398 | 1.766 | -1.066 | 5.861 | 0.175 |  |
| PC7 | -0.49 | 1.239 | -2.92 | 1.94 | 0.693 |  | 0.077 | 2.12 | -4.081 | 4.235 | 0.971 |  | -1.336 | 1.254 | -3.795 | 1.122 | 0.287 |  |
| PC8 | -1.087 | 1.216 | -3.473 | 1.298 | 0.371 |  | 0.447 | 1.985 | -3.446 | 4.339 | 0.822 |  | -6.625 | 3.444 | -13.378 | 0.129 | 0.055 |  |
| PC9 | -0.414 | 1.226 | -2.817 | 1.989 | 0.736 |  | 2.078 | 1.943 | -1.731 | 5.888 | 0.285 |  | -3.836 | 1.73 | -7.229 | -0.443 | 0.027 |  |
| PC10 | -2.666 | 1.223 | -5.063 | -0.268 | 0.029 |  | 5.549 | 2.066 | 1.498 | 9.601 | 0.007 |  | -2.903 | 1.468 | -5.783 | -0.024 | 0.048 |  |

**Table S9B. Association of BW_z_ with PRS_mother_ across cohorts.** Linear regression analyses examined the association between observed birthweight and maternal polygenic risk (PRS_maternal_) using a univariable model (Model 13). Results are presented as coefficients, beta values, standard errors (SE), 95% confidence intervals (CI), and q-values. Multiple testing was corrected using the Benjamini-Hochberg false discovery rate (FDR) method to generate q-values. POPS models were adjusted for maternal age at booking, maternal height, paternal height, and genetic principal components (PCs) 1-10, while BiB models were adjusted for maternal age at booking and PCs 1-10. **Model:** 13

**Supplementary Table 9C**

|  | **POPS** | | | | | | **BiB (EUR)** | | | | | | **BiB (SAS)** | | | | | |
| --- | --- | --- | --- | --- | --- | --- | --- | --- | --- | --- | --- | --- | --- | --- | --- | --- | --- | --- |
| Coefficient | Beta | SE | CI Lower | CI Upper | P-value | Q-value FDR | Beta | SE | CI Lower | CI Upper | P-value | Q-value FDR | Beta | SE | CI Lower | CI Upper | P-value | Q-value FDR |
| (Intercept) | -4.167 | 0.546 | -5.237 | -3.097 | 3.20e-14 |  | -0.712 | 0.095 | -0.898 | -0.525 | 1.09e-13 |  | -1.153 | 0.107 | -1.363 | -0.943 | 2.76e-26 |  |
| PRS_fetus_ | 0.245 | 0.016 | 0.213 | 0.278 | 4.90e-48 | 1.14e-47 | 0.276 | 0.02 | 0.237 | 0.316 | 1.50e-40 | 3.51e-40 | 0.193 | 0.02 | 0.153 | 0.232 | 1.10e-21 | 3.75e-21 |
| PRS_mother_ | 0.109 | 0.017 | 0.077 | 0.141 | 5.66e-11 | 6.60e-11 | 0.138 | 0.02 | 0.098 | 0.178 | 1.66e-11 | 1.93e-11 | 0.099 | 0.02 | 0.06 | 0.138 | 6.31e-0.7 | 7.36e-0.7 |
| Maternal height | 0.017 | 0.003 | 0.012 | 0.022 | 2.90e-11 |  |  |  |  |  |  |  |  |  |  |  |  |  |
| Paternal height | 0.008 | 0.002 | 0.003 | 0.012 | 8.73e-04 |  |  |  |  |  |  |  |  |  |  |  |  |  |
| Maternal age at booking | -0.007 | 0.003 | -0.013 | -0.001 | 0.032 |  | 0.026 | 0.003 | 0.02 | 0.033 | 4.60e-15 |  | 0.024 | 0.004 | 0.017 | 0.032 | 9.87e-11 |  |
| PC1 | 0.137 | 1.208 | -2.231 | 2.506 | 0.910 |  | -7.486 | 4.47 | -16.253 | 1.281 | 0.094 |  | 1.078 | 1.941 | -2.729 | 4.885 | 0.579 |  |
| PC2 | -1.193 | 1.202 | -3.551 | 1.164 | 0.321 |  | -2.423 | 4.582 | -11.409 | 6.563 | 0.597 |  | -4.013 | 1.003 | -5.981 | -2.045 | 6.56e-0.5 |  |
| PC3 | -0.149 | 1.256 | -2.613 | 2.314 | 0.905 |  | -0.562 | 3.222 | -6.881 | 5.756 | 0.861 |  | -1.22 | 1.01 | -3.201 | 0.76 | 0.227 |  |
| PC4 | 0.506 | 1.205 | -1.856 | 2.868 | 0.675 |  | -3 | 3.588 | -10.036 | 4.035 | 0.403 |  | 0.577 | 1.194 | -1.764 | 2.918 | 0.629 |  |
| PC5 | 0.623 | 1.18 | -1.691 | 2.937 | 0.597 |  | 2.506 | 2.731 | -2.85 | 7.862 | 0.359 |  | -0.304 | 1.44 | -3.127 | 2.519 | 0.833 |  |
| PC6 | 0.371 | 1.195 | -1.973 | 2.715 | 0.757 |  | -0.063 | 2.235 | -4.446 | 4.32 | 0.978 |  | 2.152 | 1.732 | -1.245 | 5.549 | 0.214 |  |
| PC7 | -0.498 | 1.187 | -2.825 | 1.83 | 0.675 |  | 1.222 | 2.033 | -2.765 | 5.21 | 0.548 |  | -1.913 | 1.229 | -4.323 | 0.497 | 0.12 |  |
| PC8 | -0.895 | 1.164 | -3.178 | 1.388 | 0.442 |  | 0.055 | 1.903 | -3.676 | 3.786 | 0.977 |  | -6.887 | 3.373 | -13.501 | -0.274 | 0.041 |  |
| PC9 | -0.601 | 1.173 | -2.902 | 1.7 | 0.608 |  | 2.634 | 1.862 | -1.017 | 6.285 | 0.157 |  | -3.356 | 1.696 | -6.681 | -0.031 | 0.048 |  |
| PC10 | -2.737 | 1.17 | -5.032 | -0.442 | 0.019 |  | 4.672 | 1.981 | 0.788 | 8.557 | 0.018 |  | -2.996 | 1.438 | -5.815 | -0.176 | 0.037 |  |
| PRS_fetus_$\times$PRS_mother_ | 0.031 | 0.016 | 0 | 0.061 | 0.048 | 0.048 | 0.007 | 0.019 | -0.031 | 0.044 | 0.718 | 0.718 | 0.036 | 0.018 | 0 | 0.071 | 0.047 | 0.047 |

**Table S9C. Association of BWz with Interactive Model in POPS and BiB.** Linear regression analyses examined the association of observed birthweight with fetal and maternal polygenic risk using an interactive model (Model 15). Results are presented as coefficients, beta values, standard errors (SE), 95% confidence intervals (CI), p-values, and q-values. Multiple testing was corrected using the Benjamini-Hochberg false discovery rate (FDR) method to generate eq0values. POPS models were adjusted for maternal age at booking, maternal height, paternal height, and genetic principal components (PCs) 1-10, while BiB models were adjusted for maternal age at booking and PCs 1-10. **Model:** 15

**Supplementary Table 9D**

|  | **POPS** | | | | | | **BiB (EUR)** | | | | | | **BiB (SAS)** | | | | | |
| --- | --- | --- | --- | --- | --- | --- | --- | --- | --- | --- | --- | --- | --- | --- | --- | --- | --- | --- |
| Coefficient | Beta | SE | CI Lower | CI Upper | P-value | Q-value FDR | Beta | SE | CI Lower | CI Upper | P-value | Q-value FDR | Beta | SE | CI Lower | CI Upper | P-value | Q-value FDR |
| (Intercept) | -4.166 | 0.546 | -5.237 | -3.096 | 3.31e-14 |  | -0.706 | 0.095 | -0.896 | -0.524 | 1.15e-13 |  | -1.157 | 0.107 | -1.367 | -0.946 | 2.08e-26 |  |
| PRS_fetus_ | 0.246 | 0.016 | 0.213 | 0.278 | 3.67e-48 | 1.14e-47 | 0.277 | 0.02 | 0.237 | 0.316 | 1.25e-40 | 3.51e-40 | 0.192 | 0.02 | 0.153 | 0.231 | 1.61e-21 | 3.75e-21 |
| PRS_mother_ | 0.11 | 0.017 | 0.078 | 0.143 | 3.58e-11 | 5.02e-11 | 0.138 | 0.02 | 0.098 | 0.178 | 1.65e-11 | 1.93e-11 | 0.1 | 0.02 | 0.061 | 0.139 | 5.05e-07 | 7.08e-07 |
| Maternal height | 0.017 | 0.003 | 0.012 | 0.022 | 3.51e-11 |  | NA | NA | NA | NA | NA |  | NA | NA | NA | NA | NA |  |
| Paternal height | 0.008 | 0.002 | 0.003 | 0.012 | 7.72e-04 |  | NA | NA | NA | NA | NA |  | NA | NA | NA | NA | NA |  |
| Maternal age at booking | -0.007 | 0.003 | -0.013 | 0 | 0.035 |  | 0.026 | 0.003 | 0.02 | 0.033 | 4.56e-15 |  | 0.025 | 0.004 | 0.017 | 0.032 | 4.64e-11 |  |
| PC1 | 0.137 | 1.208 | -2.233 | 2.506 | 0.910 |  | -7.447 | 4.272 | -16.76 | -0.004 | 0.05 |  | 1.299 | 1.939 | -2.504 | 5.103 | 0.503 |  |
| PC2 | -1.229 | 1.203 | -3.587 | 1.13 | 0.307 |  | -2.412 | 4.579 | -11.354 | 6.605 | 0.604 |  | -4.048 | 1.004 | -6.017 | -2.079 | 5.72e-05 |  |
| PC3 | -0.156 | 1.257 | -2.621 | 2.308 | 0.901 |  | -0.549 | 3.211 | -6.679 | 5.916 | 0.905 |  | -1.226 | 1.011 | -3.208 | 0.756 | 0.225 |  |
| PC4 | 0.491 | 1.205 | -1.872 | 2.854 | 0.684 |  | -2.99 | 3.523 | -10.141 | 3.676 | 0.359 |  | 0.654 | 1.194 | -1.688 | 2.995 | 0.584 |  |
| PC5 | 0.603 | 1.181 | -1.712 | 2.918 | 0.610 |  | 2.541 | 2.729 | -2.810- | 7.892 | 0.352 |  | -0.255 | 1.44 | -3.08 | 2.569 | 0.859 |  |
| PC6 | 0.5 | 1.194 | -1.842 | 2.842 | 0.676 |  | -0.085 | 2.234 | -4.466 | 4.296 | 0.97 |  | 1.974 | 1.731 | -1.421 | 5.368 | 0.254 |  |
| PC7 | -0.406 | 1.187 | -2.733 | 1.921 | 0.732 |  | 1.214 | 2.033 | -2.773 | 5.2 | 0.551 |  | -1.941 | 1.23 | -4.353 | 0.471 | 0.115 |  |
| PC8 | -0.875 | 1.165 | -3.159 | 1.41 | 0.453 |  | 0.04 | 1.902 | -3.689 | 3.77 | 0.983 |  | -6.769 | 3.374 | -13.386 | -0.152 | 0.045 |  |
| PC9 | -0.651 | 1.174 | -2.952 | 1.651 | 0.579 |  | 2.633 | 1.861 | -1.017 | 6.284 | 0.157 |  | -3.288 | 1.696 | -6.615 | 0.039 | 0.053 |  |
| PC10 | -2.691 | 1.171 | -4.987 | -0.395 | 0.022 |  | 4.679 | 1.98 | 0.796 | 8.563 | 0.018 |  | -2.981 | 1.439 | -5.803 | -0.16 | 0.038 |  |

**Table S9D: Associations between PRS_fetus_, PRS_mother_, and BW_z_ across POPS, BiB-EUR, and BiB-SAS with all predictors.** We assessed the cross-sectional associations of PRS_mother_ and PRS_fetus_ using the joint model across cohorts. We reported regression coefficients (β), standard errors (SE), 95% confidence intervals (CI), p-values, and q-values. Adjustments in POPS included maternal age at booking, maternal height, paternal height, PCs 1-10, while BiB adjusted for maternal age at booking and PCs 1-10. False discovery rate-adjusted p-values were calculated using the Benjamini-Hochberg method to generate q-values. This table corresponds to Table 2 in the main text, which shows only the main predictors of interest. **Model:** 14

**Supplementary Figures 3A,B&C**

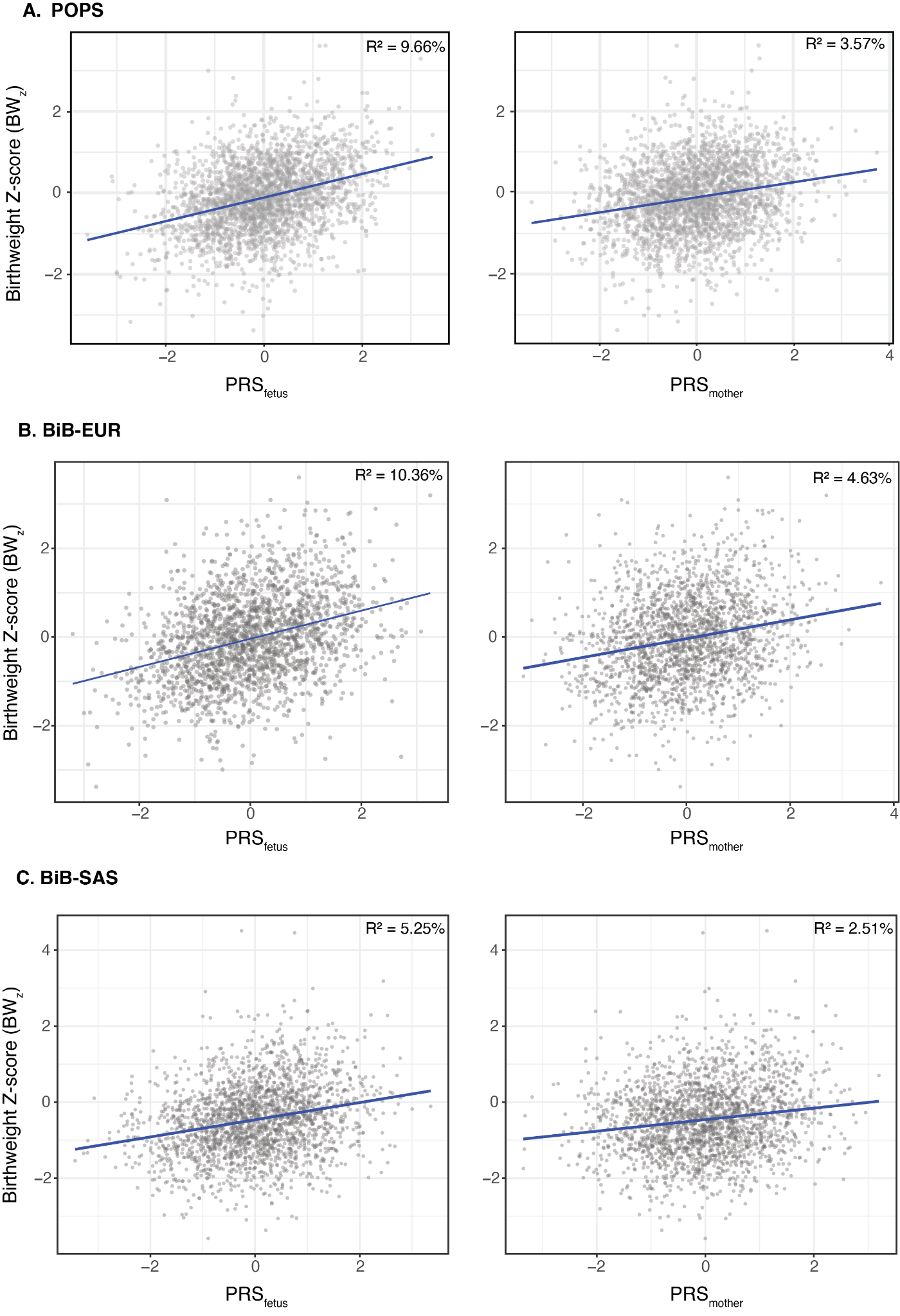

**Figures S3A,B&C. Association between fetal and maternal birthweight polygenic risk scores (PRS) and observed birthweight Z-score (BW_z_)**. Associations were estimated from a regression of BW_z_ on each PRS separately, adjusted for covariates (model 12-13, Methods). The left panel shows the association with fetal BW PRS (PRS_fetus_) and the right panel shows the association with maternal birthweight PRS (PRS_mother_). R^2^ is the incremental variance explained by each PRS over covariates alone. BiB-EUR:genetically-inferred European ancestry participants from Born in Bradford (N=2,075); BiB-SAS: genetically-inferred South Asian ancestry from Born in Bradford (N=2,210); POPS: Pregnancy outcomes prediction study (N=2,537).

**Supplementary Table 10A**

|  | **POPS** |  |  | **BiB (EUR)** |  |  | **BiB (SAS)** |  |  |
| --- | --- | --- | --- | --- | --- | --- | --- | --- | --- |
| Model | R^2^ | ΔR^2^ | AIC | R^2^ | ΔR^2^ | AIC | R^2^ | ΔR^2^ | AIC |
| Model 11: Base (covariates only) | 5.03% |  | 6113.8 | 3.61% | NA | 5717.1 | 4.14% | NA | 5950.7 |
| Model 12: Fetal PRS | 14.69% | 9.66% | 5851.6 | 13.97% | 10.36% | 5483.1 | 9.39% | 5.25% | 5828.1 |
| Model 13: Maternal PRS | 8.60% | 3.57% | 6021.5 | 8.25% | 4.63% | 5616.8 | 6.64% | 2.51% | 5894.1 |
| Model 14: Fetal + Maternal PRS (Joint) | 16.21% | 11.18% | 5809.5 | 15.85% | 12.23% | 5439.5 | 10.43% | 6.29% | 5804.7 |
| Model 15: Fetal × Maternal (Interactive) | 16.34% | 11.31% | 5807.6 | 15.85% | 12.24% | 5441.1 | 10.59% | 6.45% | 5802.7 |

**Table S10A. Comparison of model performance for predicting BW_z_ in POPS and BiB. In POPS and** BiB, we generated R^2^ and AIC values to represent the variance in birthweight explained by the covariate-only (Model 11), fetal (Model 12), maternal (Model 13), joint (Model 14), and interactive (Model 15) linear models. All ΔR^2^ values were calculated in reference to the covariate-only model. Adjustments for POPS included maternal age at booking, maternal height, paternal height, PCs 1-10. Adjustments for BiB included maternal age at booking and PCs 1-10. **Models:** 11-15

**Supplementary Table 10B**

|  | **POPS** | **BiB (EUR)** | **BiB (SAS)** |
| --- | --- | --- | --- |
| Genetic Component | Variance explained % | Variance explained % | Variance explained % |
| Independent fetal effect (ΔR^2^ fetal PRS) | 7.61 | 7.6 | 3.78 |
| Independent maternal effect (ΔR^2^ maternal PRS) | 1.52 | 1.87 | 1.04 |

**Table S10B. Variance decomposition for BW_z_ in POPS and BiB.** We calculated the proportion of variance in BW_z_ explained independently by PRS_fetus_ and PRS_mother_. Independent fetal and maternal effects were calculated by subtracting the variance explained by the fetal (Model 2) and maternal (Model 3) models from that explained by the joint (Model 4) model. Adjustments for POPS included maternal height, paternal height, maternal age at booking, PCs 1-10. Adjustments for BiB included maternal age at booking and PCs 1-10. **Models:** 12-14

**Supplementary Figure 4**
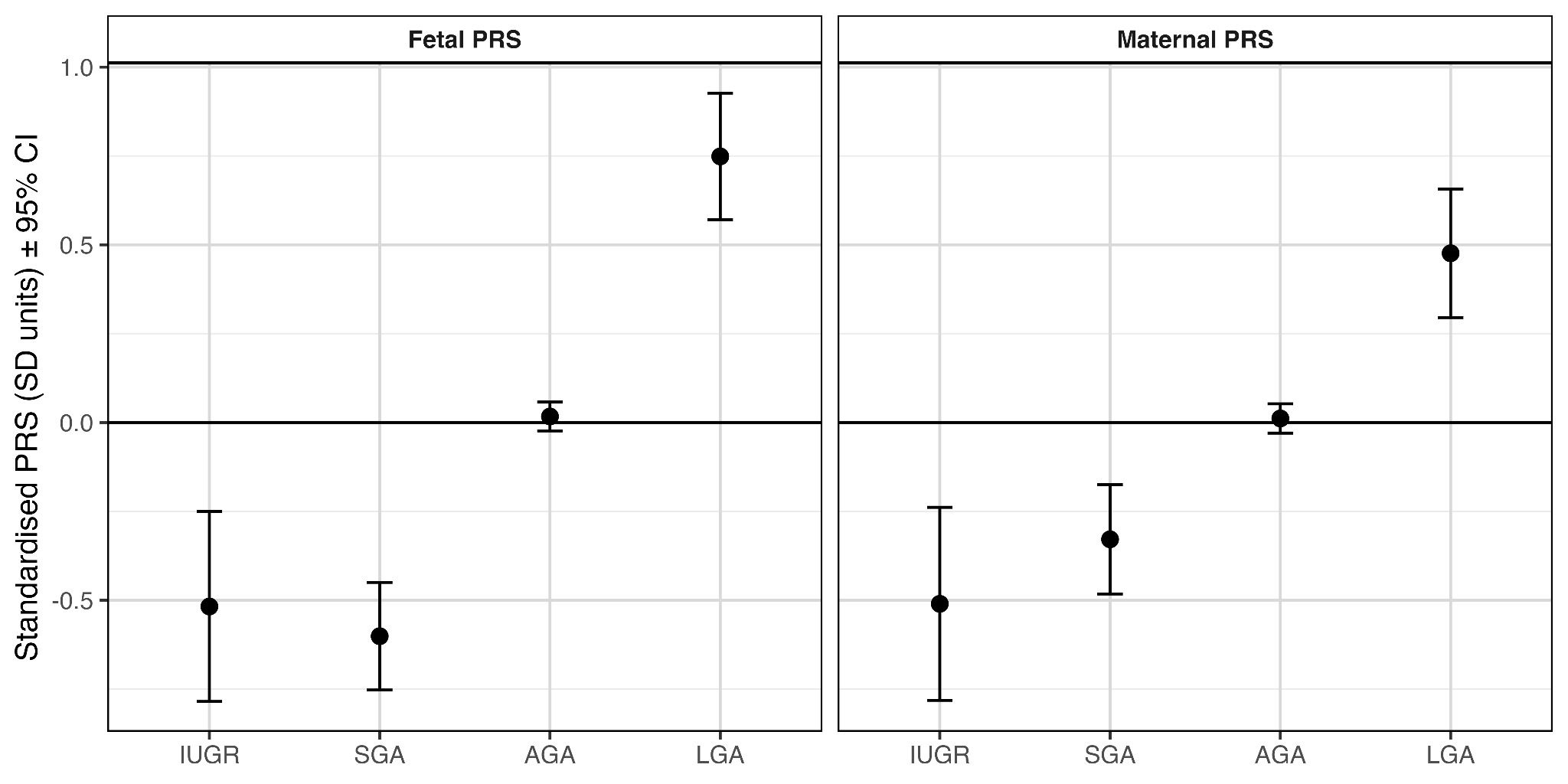

**Figure S4. Mean Residualized and Standardized PRS_fetus_ and PRS_mother_ across Birthweight Classifications in POPS.** Fetal and maternal plots for POPs show residualized and standardized mean PRS_fetus_ and PRS_mother_ for each birthweight classification with 95% confidence intervals. MoBa does not have an IUGR phenotype so mean PRS and CIs for SGA, AGA, and LGA are reported.

**Supplementary Table 11A**

| **Growth category** | **N** | **PRS_fetus_ mean** | **PRS_fetus_ SD** | **PRS_mother_ mean** | **PRS_mother_ SD** |
| --- | --- | --- | --- | --- | --- |
| IUGR | 51 | -0.518 | 0.829 | -0.510 | 0.989 |
| SGA | 159 | -0.601 | 1.044 | -0.329 | 0.964 |
| AGA | 2,218 | 0.017 | 0.970 | 0.011 | 0.993 |
| LGA | 115 | 0.749 | 0.995 | 0.476 | 0.946 |

**Table S11A. Summary of residualized PRSs across birthweight group.** In POPS, PRSs were residualized on the first 10 genetic principal components (PCs) prior to analysis. Mean and standard deviation (SD) values were calculated for both maternal and fetal polygenic risk scores within each birthweight group.

**Supplementary Table 11B**

| **Contrast** | **Mean Difference** | **SE** | **df** | **Lower CI** | **Upper CI** | **P-value** | **Significance** |
| --- | --- | --- | --- | --- | --- | --- | --- |
| SGA - AGA | -0.618 | 0.085 | 178.102 | -0.787 | -0.450 | 7.5e-11 | *** |
| LGA - AGA | 0.732 | 0.095 | 125.501 | -0.920 | -0.544 | 2.1e-11 | *** |
| SGA - LGA | -1.350 | 0.124 | 252.437 | -1.595 | -1.105 | 5.2e-22 | *** |
| IUGR - SGA | 0.084 | 0.143 | 105.201 | -0.199 | 0.366 | 1.000 |  |
| IUGR - AGA | -0.535 | 0.118 | 53.195 | -0.771 | -0.298 | 2.0e-04 | *** |
| IUGR - LGA | -1.266 | 0.149 | 113.858 | -1.561 | -0.972 | 4.5e-13 | *** |

**Table S11B. Pairwise t-testing of PRS_fetus_ between birthweight classifications.** We residualized PRS_mother_ and PRS_fetus_ on principal components, standardized the residualized PRS values, and calculated mean PRS_mother_ and PRS_fetus_ for each birthweight classification. We conducted pairwise t-tests to evaluate differences in mean polygenic risk between birthweight groups. The second group in each comparison is the reference group. We reported the mean PRS difference, standard error, DF, 95% CIs, and P-values. The Bonferroni correction was applied to all p values.

**Supplementary Table S11C**

| **Contrast** | **Mean Difference** | **SE** | **df** | **Lower CI** | **Upper CI** | **P-value** | **Significance** |
| --- | --- | --- | --- | --- | --- | --- | --- |
| SGA - AGA | -0.340 | 0.079 | 182.892 | -0.497 | -0.184 | 1.7e-04 | *** |
| LGA - AGA | 0.465 | 0.091 | 127.382 | -0.644 | -0.285 | 6.4e-06 | *** |
| SGA - LGA | -0.805 | 0.117 | 248.417 | -1.035 | -0.575 | 2.6e-10 | *** |
| IUGR - SGA | -0.182 | 0.158 | 82.644 | -0.496 | 0.133 | 1.000 |  |
| IUGR - AGA | -0.522 | 0.140 | 52.343 | -0.803 | -0.240 | 0.003 | ** |
| IUGR - LGA | -0.986 | 0.164 | 92.111 | -1.313 | -0.660 | 2.2e-07 | *** |

**Table S11C. Pairwise t-testing of PRS_mother_ between birthweight classifications.** We residualized PRS_mother_ and PRS_fetus_ on principal components, standardized the residualized PRS values, and calculated mean PRS_mother_ and PRS_fetus_ for each birthweight classification. We conducted pairwise t-tests to evaluate differences in mean polygenic risk between birthweight groups. The second group in each comparison is the reference group. We reported the mean PRS difference, standard error, DF, 95% CIs, and P-values. The Bonferroni correction was applied to all p values.

**Supplementary Table 12**

| **Birthweight category** | **N** | **Mean D** | **Median D** | **SD D** | **IQR D** |
| --- | --- | --- | --- | --- | --- |
| IUGR | 51 | -1.624 | -1.590 | 0.565 | 0.764 |
| SGA | 159 | -1.331 | -1.289 | 0.502 | 0.581 |
| AGA | 2,218 | 0.050 | 0.034 | 0.615 | 0.876 |
| LGA | 115 | 1.592 | 1.510 | 0.557 | 0.674 |

**Table S12. Distribution of deviation from genetically predicted birthweight across fetal growth categories in the POPS cohort.** Summary statistics of deviation (D = observed minus genetically predicted birthweight) stratified by fetal growth classification in the POPS cohort. For each group, we report sample size (N), mean and median deviation, standard deviation (SD), and interquartile range (IQR). Negative values indicate birthweights lower than genetically predicted, while positive values indicate higher-than-predicted birthweights.
