## Supplemental File 2 for "Contribution of maternal and fetal genetics to fetal weight throughout pregnancy and at birth"

### Supplemental File 2: Processing and Preparation of Genotype Data in the Pregnancy Outcome Prediction Study (POPS) and Born in Bradford (BiB)

a. **Pregnancy Outcome Prediction Study (POPS)**

POPS maternal blood and umbilical cord samples were genotyped using the Illumina Infinium Global Screening Array Kit (GSA v3). 7,890 samples (4,048 maternal DNA samples and 3,854 cord DNA samples) were analyzed; 654,027 variants were directly genotyped. 258 samples were excluded due to genotyping call rates >95% and 197 due to high/low heterozygosity rate^1^. We estimated kinship using the Kinship-based Inference for Genome-wide association studies algorithm (KING) based on 636,695 directly genotyped SNPs and excluded 10 confirmed genetic duplicates. Imputation was performed with Minimac4 on the 7,425 samples remaining using the NHLBI TOPMed reference panel. After imputation, we excluded 48 parent-offspring pairs (n=96) that did not have a first-degree relationship (kinship estimation <0.177)^2^; full siblings and participants with a second-degree or closer relationship with another participant outside their mother-child pair were also excluded. We conducted a subsequent kinship analysis on third degree relatives and excluded 64 children and 48 mothers, resulting in a fetal sample of 2,825 and a maternal sample of 2,585. The complete, matched analytical sample included 2,537 mother-child pairs.

Principal component analysis (PCA) was performed to evaluate population structure using PLINK v2.00 alpha (n=50,617 variants). Genotype data were restricted to typed variants and filtered to retain common variants (minor allele frequency > 0.05). Linkage disequilibrium (LD) pruning was conducted using a sliding window of 50 variants, advancing by 5 variants per step, removing variant pairs with r^2^ > 0.1. The resulting LD-pruned dataset was used for PCA. Principal components were computed using PLINK’s approximate PCA implementation; the top 10 components were retained for downstream analyses.

b. **Born in Bradford (BiB)**

BiB samples were genotyped using two chips, the Infinium CoreExome-24 v.1.1 BeadChip (~550 K SNPs), and (2) the Infinium GSA-24 v.1 (~640 K SNPs). We merged the two batches, and initial quality control included exclusion of variants with missingness >1% and individuals with missingness >10%, and removal of duplicated variants. We further excluded 110 samples with sex discrepancies. Relatedness inference was performed using KING, and we removed 123 genetic duplicates, 465 individuals who lacked an expected first-degree relationship, and five individuals with an unexpected inferred first-degree relationship.

Genetic ancestry was inferred using principal component analysis (PCA). PCA was performed on a subset of autosomal, biallelic, SNPs with minor allele frequency (MAF) >5%, missingness <1%, and passing Hardy–Weinberg equilibrium (P>1×10⁻²⁰). SNPs in 24 long-range LD regions were excluded and LD pruning (pairwise r2 <0.2 in batches of 50 SNPs with sliding windows of 5) was performed. PCA was conducted in unrelated samples from the 1000 Genomes Project (1kG, build 37) using a subset of overlapping SNPs (N=63,015), and all BiB samples were projected into this principal component (PC) space. Genetic ancestry was assigned using a support vector machine classifier applied to the PCs, and individuals with discordant self-reported and genetically-inferred ancestry were excluded (N=220). PCA was then repeated within each ancestry group in BiB using a subset of unrelated individuals following additional variant-level QC and LD pruning. All remaining BiB samples were projected into the resulting ancestry-specific PC space, and loadings for the first ten principal components were calculated separately within each ancestry group.

Genotype imputation was performed separately for each batch of genotype array data using the Michigan Imputation Server^3^ with the GenomeAsia Pilot (GAsP) reference panel^4^ for SAS participants and the Haplotype Reference Consortium panel for EUR participants^5^. After imputation, high-quality imputed SNPs were retained (Minimac4 R^2^ > 0.8 and MAF > 1%), and GSA and CoreExome datasets were merged. Variants with >1% missingness in the combined dataset were excluded, leaving 6,608,017 and 3,525,222 variants for analysis in the EUR and SAS ancestry groups, respectively.

For downstream analyses, we restricted to a subset of unrelated children (kinship < 0.0442; no closer than third-degree relatives) identified from LD-pruned genotype data.
